# Personalized Machine Learning guided Intervention for Optimizing Lifestyle Behaviors in Depression

**DOI:** 10.1101/2025.11.04.25339502

**Authors:** Jason Nan, Suzanna Purpura, Satish Jaiswal, Houtan Afshar, Vojislav Maric, James K. Manchanda, Charles T. Taylor, Dhakshin Ramanathan, Jyoti Mishra

## Abstract

**Objective:** Personalized data-driven interventions for depression are much needed. Here, we leveraged N-of-1 machine learning (ML) to optimally target behavioral lifestyle interventions for depression.

**Methods:** 50 individuals with mild-to-moderate depression enrolled in the single-arm, open-label Personalized Mood Augmentation (PerMA) clinical trial (NCT05662254). Participants completed a two-week digital monitoring phase using smartphone-based ecological momentary assessments (EMAs, 4x/day) plus smartwatch tracking of mood and lifestyle factors (sleep/exercise/diet/social connection). Personalized ML models were generated from these data to identify lifestyle factors most predictive of individual mood, and results were translated to individualized mood augmentation plans (iMAPs) implemented by participants for six weeks with once-a-week health coach guidance.

**Results:** Intervention completers (n=40) showed significant reduction in depression symptoms (primary outcome self-rated PHQ9 −3.5±3.8, Cohen’s d=-0.89, CI [−1.25 −0.53], p<0.001; clinician-rated HDRS -7.2±6.8, d=-1.03, CI [−1.41 −0.65], p<1E-6) with benefits sustained up to 12-week follow-up. Co-morbid anxiety was also significantly reduced (GAD7: d=-0.85, CI [−1.2, −0.49], p<0.001) and quality of life improved (d=0.68, CI [0.33, 1.02], p<0.001). Additionally, objective cognitive measures impacted in depression including selective attention (d=0.51, CI [0.18, 0.84], p<0.001), interference processing (d=0.53, CI [0.2, 0.85], p<0.01) and working memory (d=0.66, CI [0.31, 0.99], p<0.001) showed significant enhancement. EMA tracking confirmed that improvement in depressed mood was specifically predicted by improvement in individually targeted lifestyles (β=0.4±0.09, p<0.0005).

**Conclusion:** The PerMA trial presents a robust personalized lifestyle intervention approach for depression and merits scale-up and RCT testing.

## Introduction

Depression carries the largest burden of mental health disorders with prevalence of depressive episodes observed in 18% of the total US population and 21% of young adults ^1,2^. Besides the negative health impacts of depression, its socio-economic cost burden was recently estimated at greater than $380 billion with healthcare as the major cost driver alongside household and work-related costs ^3^. Notably, the majority (∼67%) of all depression cases fall in the category of mild-to-moderate depression ^3^ for which integrative behavioral health treatments are recommended first-line as effective and scalable solutions ^4^. These include physical activity/exercise, dietary modification, adequate sleep and social interaction, as well as mindfulness-based meditation, all of which have shown treatment efficacy in separate clinical trials of depression ^5–17^. Meta-analyses of lifestyle intervention trials find low-to-medium effect sizes for depression alleviation (Cohen’s d∼0.3-0.5). Yet, a major limitation of these homogenously assigned (non-personalized) behavioral intervention studies is the heterogeneity across individuals of the specific lifestyle factors most closely linked to their depression. Consequently, a single lifestyle treatment domain is unlikely to be universally beneficial, and these studies fail to account for individual lifestyle differences that may favor one behavioral health solution over another ^18–20^. This limitation has sparked a new push towards developing personalized treatments that are tailored to each individual. Personalized behavioral health solutions for mild-to-moderate depression could prove to be more effective, scalable and accessible, hence, optimizing such treatments offers incredible benefit for society.

Studies across multiple fields of healthcare have shown that personalizing treatment leads to higher treatment adherence as well as improved satisfaction ^21–24^. A depression intervention study that used a personalized advantage index (PAI) to assign either cognitive or interpersonal therapy to individuals showed decreased long-term symptom severity in patients receiving PAI-indicated vs. PAI-non-indicated treatment ^25^. A recent systematic review and meta-analysis of multiple personalized psychological interventions found that personalization significantly improves the effect size of treatment compared to standard therapy ^26^. Yet, of note, all of these personalized intervention studies use predictive models based on multi-subject data. While this may be adequate to distinguish between subtypes of depression, such models do not capture individual lifestyle attributes that drive day-to-day fluctuations in depressed mood. Further, such models based on data from specific populations may not be generalizable and thus may fail to properly assign the optimal treatment ^27^. An alternate approach to population data-based machine learning is to design models that link lifestyle features with depressed mood based on an individual’s own data over time – also referred to as idiographic or N-of-1 modeling. Here, based on our prior research developing an N-of-1 machine learning (ML) pipeline for depressed mood ^18^, we aimed to assign behavioral interventions exclusively based on individual data acquired over time and thereby, steer clear of issues related to between-subject heterogeneity and generalizability.

Overall, this study aimed to deliver personalized behavioral intervention based on data-driven N- of-1 ML models tailored to individual lifestyles. We have previously demonstrated high accuracy ML modeling of individual depressed mood based on ecological momentary assessments (EMA) and passive smart watch data ^18,19,28^. Furthermore, leveraging model explainers such as Shapley plots, we have identified the top predictive intervention targets for each individual ^18,19^. Trained health coaches can then utilize this predictive information to determine the optimal behavioral intervention for each individual. Delivering behavioral health interventions through health coaches has the added benefit of being cost-effective compared to repeat visits to licensed mental healthcare providers, i.e., psychiatrists or clinical psychologists, and can be particularly beneficial in low-income settings ^29–31^. Additionally, we used a multidimensional approach to evaluate trial outcomes; symptom assessments were complemented with evaluation of change in subjective mental health behaviors and quality of life, as well as change in objective measures of neuro- cognition that are impacted in depression ^32–38^. Overall, to the best of our knowledge, this is a first study to implement a N-of-1 ML-personalized behavior intervention for depressed mood using digital lifestyle markers. If shown to be effective, such a personalized approach can be a promising way to remotely deliver scalable treatment for depression that is optimized to individual lifestyles.

## Methods

### Participants

50 individuals participated in the study (mean age: 43.3 ± 16.1, range: 23-76, 12 males). Participants were recruited from the San Diego community using the ResearchMatch registry and social media, email and print flyers. All participants were fluent in English and study inclusion was based on current mild-to-moderate depression symptoms per self-reported Patient Health Questionnaire (PHQ9) scores in the 5-17 range^39^ further confirmed via structured clinical interview for DSM V Axis I disorders (SCID) ^40^. Exclusion criteria evaluated via SCID consisted of evidence for active substance use disorder, psychotic disorder, bipolar disorder, eating disorder, or displaying acutely suicidal behaviors. Participants aged 60 years or older completed the Montreal Cognitive Assessment (MoCA ≥ 26) ^41^. All participants provided written informed consent in accordance with the Declaration of Helsinki before participating in the study. All experimental procedures were approved by the Institutional Review Board of the University of California San Diego (UCSD) (protocol #180140). Data collection took place during Winter 2022- Fall 2024.

### Sample Size and Power

The sample size of 50 participants accounted for up to 20% attrition, so as to obtain at least 40 complete study samples that were powered to detect a statistically meaningful medium effect size (Cohen’s d>0.45) primary outcome difference in pre vs post comparisons of PHQ9 scores at 0.8 power and alpha level of 0.05, computed a priori ^42^.

### Study Procedure

We pre-registered the single-arm study in ClinicalTrials.gov as the Personalized Mood Augmentation (PerMA) trial: NCT05662254 ^43^.

During the first phase, participants downloaded the Unity-based BrainE© app on their iOS/Android smartphone to complete daily EMAs of mood and lifestyle factors up to 4 times per day until they completed 60 sessions (2-4 weeks). ^44^. The app sent regular notifications daily to all participants following the methodology of recent research on longitudinal mood monitoring ^18,19,45^. Participants also wore a Samsung Galaxy wristwatch throughout the study. At completion of 60 sessions of mood and lifestyle tracking, individual mood augmentation plans (iMAPs) were generated using N-of-1 machine learning models ^18,19^, targeting one of four major lifestyle domains: sleep, exercise, diet, and social connection (see Supplementary Methods).

The second phase of the study lasted 6 weeks where participants had once-a-week iMAP guided sessions (GS) with a trained health coach on one-on-one virtual video calls of ∼20 minutes duration each. Health coaches in this study were two health sciences trainees (co-authors HA, VM) who received basic training on how to read and interpret iMAPs in layman terms, and then how to motivate and guide individuals through standard, evidence-based behavioral lifestyle interventions focused on sleep, exercise, diet or social connection – protocols described in the Supplementary Methods. The coaches received about 8-10 hours of training consisting of both didactic training as well as practical skills development with role play of realistic participant-coach interactions supervised with feedback from the study psychiatrist (co-author DR). The study psychiatrist also monitored the first few video GS performed by each of the newly trained health coaches to ensure coaching consistency and provide any additional coaching feedback, if necessary. During the 6- weeks iMAP implementation phase, participants continued to complete once-a-day mood and lifestyle EMAs to track daily progress. **Figure 1** visualizes the study flow and protocol design.

**Figure 1.**
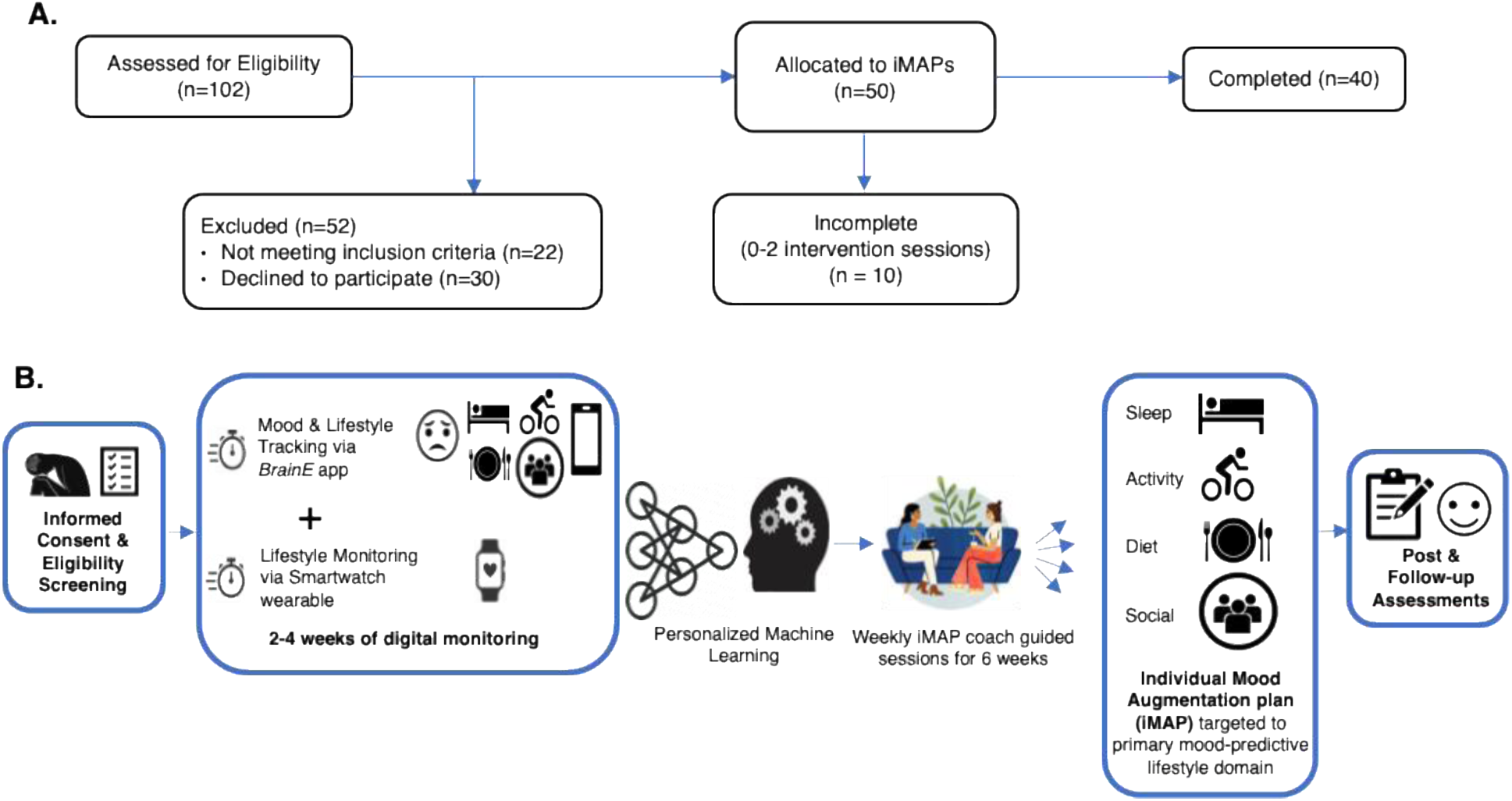
Personalized Mood Augmentation (PerMA) trial flow. (A) Participant flow is shown from eligibility assessments to completion. (B) Study design is shown from left to right. Informed consent and eligibility screening was followed by a 2-4 week digital monitoring phase with both smartphone and smartwatch tracking until 60 EMA sessions were completed followed by personalized machine learning model generation, which was then followed by the second phase of individual mood augmentation plan (iMAP) assignment and weekly guided sessions with a health coach for 6 weeks, and subsequent post-intervention assessments and then follow-up assessments at 6 and 12 weeks after end of intervention.

Once all the iMAPs were assigned and reviewed by DR, an automated plan assignment model was created using LLM and mathematical models and validated based off this initial dataset (see Supplementary Methods & Results).

### Clinical and Behavioral Outcomes

The primary clinical outcome was change in self-reported depression ratings on the PHQ9 scale ^39^ from pre- to post-intervention. The PHQ9 scale was completed by self-report at eligibility screening; before start of study digital monitoring procedures (pre); at end of digital monitoring/beginning of the second phase at the first coach guided session (GS1); every two weeks during iMAP intervention (at GS3, GS5); at post-intervention (post); and also at 6-week and 12-weeks follow-up after end of intervention.

Secondary clinical and behavioral outcomes included the Generalized Anxiety Disorder 7-item scale (GAD7 ^46^) acquired at the same time-points as the PHQ9 above, given that anxiety and depression are highly co-morbid ^47,48^. In addition, at pre- and post-intervention time-points, outcome scores were acquired for clinician-rated depression on the Hamilton Depression Rating Scale (HDRS ^49^), self-rated quality of life reported on the Short Form 12 Mental Component Summary (MCS12 ^50^), and mindfulness self-rated per the Mindful Attention Awareness Scale (MAAS ^51^). Mindfulness was intentionally assessed as it can facilitate behavior change through greater self-awareness and control over decision-making ^52,53^.

All clinical and behavioral assessments were completed in a de-identified manner with alphanumeric study IDs using the secure web-based Ready Electronic Data Capture (REDCap) system.

### Cognitive Outcomes

Secondary outcomes also included assessments of neuro-cognition completed using the BrainE app, specifically tests of selective attention, interference processing, working memory and emotion bias, described in several prior publications ^18,54–66^ and also in Supplementary Methods.

### Data Analyses

For all analyses, we report effect sizes as Cohen’s d for post (or follow-up) vs. pre-intervention comparisons with 95% confidence intervals (CI). For regression analyses, standardized regression coefficients are reported where applicable: >0.1 as small, >0.3 as medium, and >0.5 as large effect size ^67^.

#### Clinical and Behavioral Outcomes Analysis

Depression (PHQ9, primary outcome) and anxiety (GAD7, secondary outcome) were tracked at multiple time points - screening (PHQ9 only), pre-intervention, end of intervention phase 1/GS1, GS3, GS5, post, 6-week and 12-week follow-ups.

We ran Friedman’s repeated measures test to investigate significant changes from screening to post-intervention. In addition, we ran signed rank tests for individual time points against the ‘Pre’ time point. Secondary metrics were assessed between pre and post time points using sign rank or paired ttests when appropriate. Additionally, ANOVA was used to verify if iMAP intervention domain was a significant covariate of post vs. pre change for these outcomes. Lastly, we investigated whether any baseline metrics would be indicators of remission, which can be found in the Supplementary Methods and Results.

#### Cognitive Outcomes Analysis

For all secondary outcomes of cognition, i.e., selective attention, interference processing, working memory and emotion bias, task efficiency as the product of accuracy and processing speed was evaluated as the main outcome measure.

EMA Analysis (during Intervention Phase 2). This analysis aimed to investigate whether targeted change in the participant’s iMAP lifestyle domain versus non-specific general lifestyle change was associated with improvement in depressed mood in Intervention Phase 2. A linear fit was applied to each participant’s target and off-target domain’s representative data to calculate the slope of change over the intervention period. A robust linear regression model was fit to investigate whether slope of change of depressed mood was predicted by the change slopes for the target and off-target metrics across all participants. Data were normalized across subjects for this linear regression to report standardized betas reflecting effect size. Details about how the representative data was calculated can be found in the Supplementary Methods.

## Results

Average age of participants was 43±16 years and majority of study participants were female, per known greater prevalence of depression in females. Ethnicity data matched percentages of ethnic populations in the US San Diego county where the study was conducted. Average baseline depression and anxiety scores were in the mild-to-moderate range and were significantly correlated (rho = 0.61, p<0.0001). Full demographic data is presented in Supplementary Table 1.

### Intervention feasibility and adherence

All 50 enrolled participants completed pre and post surveys as well as the digital monitoring - phase 1 of intervention. Across all participants, the best fit personalized ML model showed high accuracy of 75.3 ± 15.2% (100 – mean ± sd of best-fit model mean absolute percentage error, MAPE). ML model mean and sd accuracies stratified by iMAP domains and iMAP distribution are shown in Figure 2; example Shapley plots of four participants assigned to one of each iMAP domain are also shown. 80% participants (n = 40 of 50) also completed intervention phase 2 with coach guided sessions, completing 19.9 ± 13 (mean ± sd) EMA over the 6 weeks of phase 2. 10 participants did not complete phase 2, i.e., 0 GS (n = 4), 1 GS (n = 4), 2 GS (n =1), or 3 GS (n =1). There was no significant difference in baseline demographics or depression scores between trial completers vs. non-completers (p>0.8).

**Figure 2.**
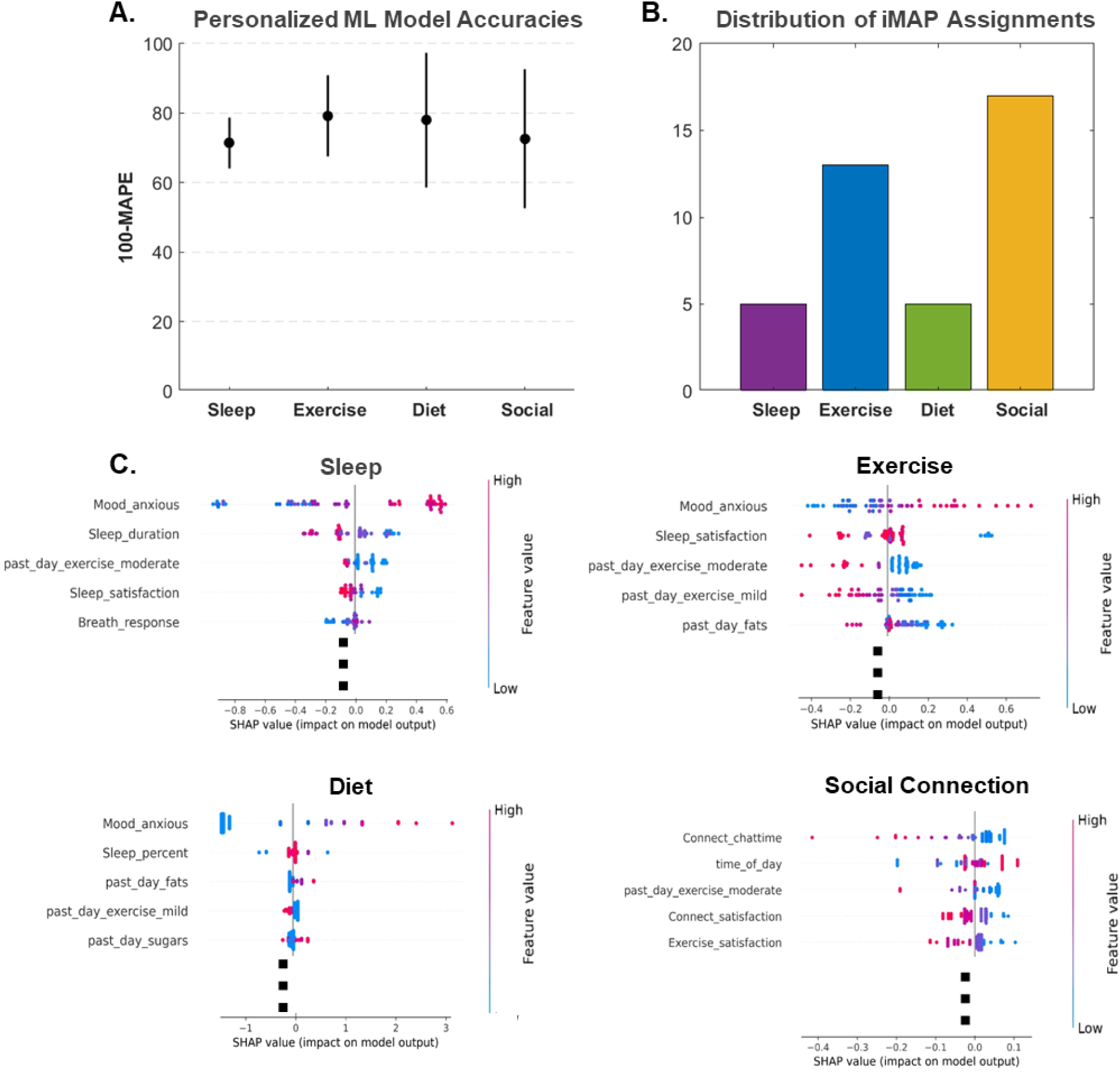
Results of personalized ML modeling. (A) ML Model accuracies split by the 4 assigned iMAP domains calculated as (100 – mean ± sd of best-fit model mean absolute percentage error, MAPE). ML model accuracies were not significantly different between domains as confirmed by one-way ANOVA (p>0.66). (B) Distribution of iMAPs assigned to the 40 study completers: sleep (5), exercise (13), diet (5), and social connection (17). (C) Four example Shapley plots showing top 5 predictive features of depressed mood from four distinct participants that were recommended either sleep, exercise, diet or social connection based iMAPs. Shapley dot plots of all ranked feature predictors show directionality of prediction; each dot represents a single datapoint, red dots indicate larger positive feature values while blue dots indicate larger negative feature values. X axis is the Shapley values with center point at 0. Positive Shapley values increase the model output (higher depression), negative Shapley values decrease the model output (lower depression). Each plots has multiple iMAP domain-specific variables appear high in the rankings, and directionality aligned with the goals of the behavioral intervention (i.e., better sleep, more exercise, lower fats and sugars, and more social connection are linked to lower depression).

### Improvements in primary depression outcome

Figure 3A shows the progression of self-reported depression symptom scores (PHQ9) across all 40 trial completers as well as separately for the 10 non-completers. The Friedman’s repeated measures ANOVA showed a significant decline in depression from pre to post-intervention (mean ± sd change = −3.5±3.8, 𝜒^2^ (df=5) = 72.6, p<1E- 13, effect size d = −0.89, CI [−1.25, −0.52]), with post-hoc testing showing significant decline from pre-intervention at all GS 1, 3, 5 and post-intervention time points. While PHQ9 scores significantly improved even after phase 1, (GS1), end of phase 2 results were significantly better than end of phase 1 results (signed rank z-value = 3.9, p=1E-4). Overall, 22 of 40 (55%) participants showed symptom remission (PHQ9 score<5) at post-intervention. Notably, PHQ9 symptom improvement was sustained at 6-week (d = −1.06, CI [−1.51, −0.6], p<1E-5, n=28) and 12-week follow-ups (d = −0.52, CI [-0.95, −0.09], p<0.05, n=22). Supplementary figure 4 shows the similar progression of PHQ9 scores for trial completers split by the 4 iMAP target lifestyle domains.

**Figure 3.**
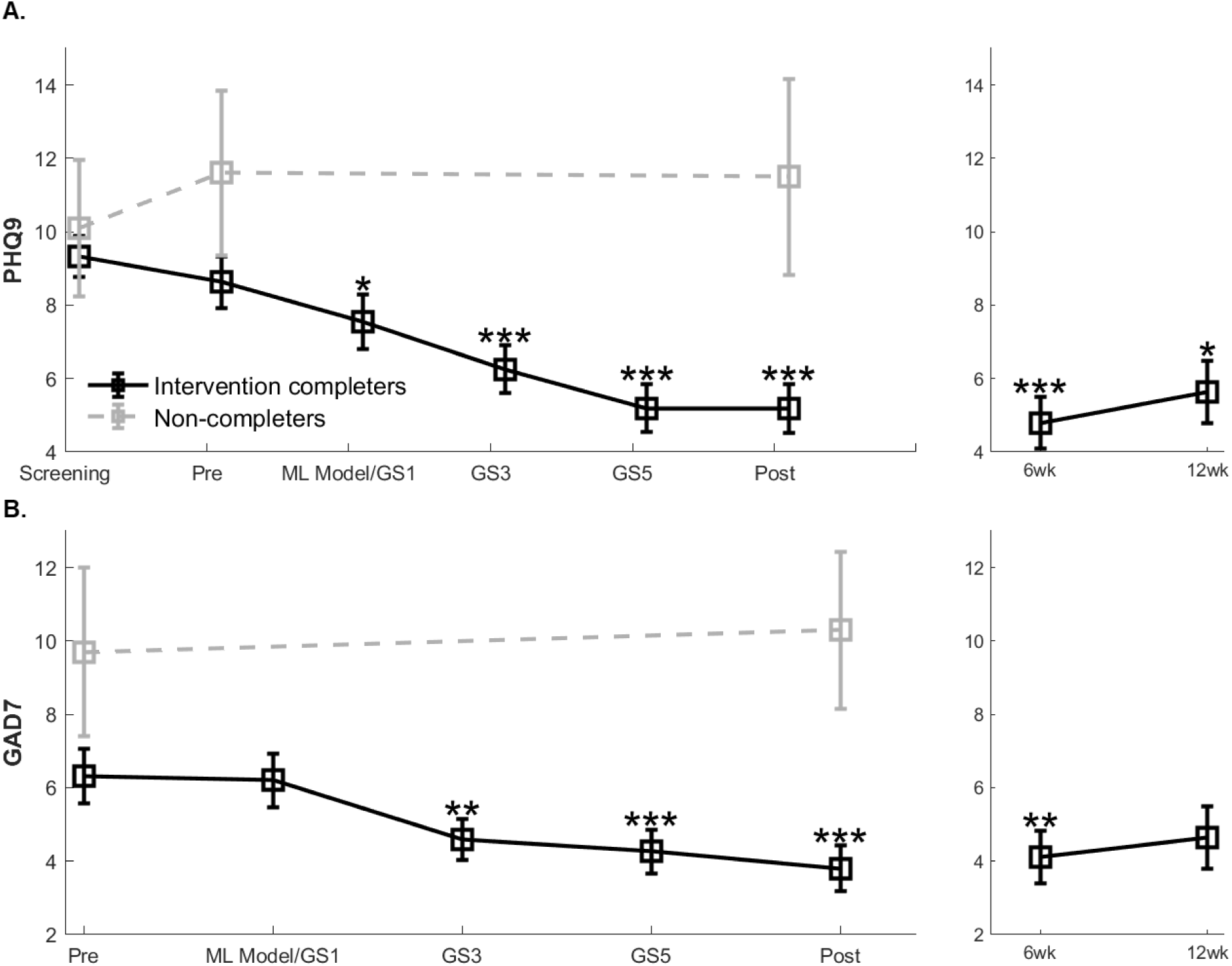
Depression (PHQ9) and anxiety (GAD7) outcomes throughout the study period. Mean ± sem scores are plotted in black for trial completers (n=40) and as dashed grey lines for non-completers (n=10). (A) PHQ9 scores are shown across the study period from screening to post-intervention with follow-up scores shown at right at 6 weeks (n=28) and 12 weeks (n=22). (B) GAD7 scores are shown across the study period from pre to post-intervention with follow-ups at right. GS: guided session with GS1 corresponding to the first coach meeting to discuss the ML model results and iMAP with each participant. Stars indicate fdr-corrected signed rank significance across all time points relative to pre-intervention: *: p<0.05; **p<0.01; ***p<0.001.

### Improvements in secondary outcomes

Figure 3B shows the progression of self-reported anxiety symptoms (GAD7) for trial completers (n=40) and separately for the non-completers (n=10). Similar to change in depression scores, the Friedman’s repeated measures ANOVA showed a significant decline in anxiety from pre to post-intervention (mean ± sd change = -2.5±3, 𝜒^2^ (df=4) = 31.9; p<1E-5, d = −0.85, CI [−1.21, −0.49]), with post-hoc testing showing significant decline from pre-intervention at GS 3, 5 and post-intervention time points, which was sustained at 6-week (d = −0.59, CI [-0.98, −0.19], p<0.005, n=28) but not 12-week follow-up (d = −0.3, CI [-0.71, 0.12], p=0.16, n=22).

Additionally, there was no effect of iMAP domain covariate (sleep/exercise/diet/social connection) in the pre- to post-intervention repeated measures ANOVA for depression (p=0.78) or anxiety (p=0.19). Demographics of age, gender, ethnicity also did not influence these outcomes (p>0.27). Notably, the 10 study non-completers did not show significant change in PHQ9 scores at post- vs. pre-intervention (d = −0.01, CI [-0.85, 0.83], p>0.8) nor GAD7 (d = 0.08, CI [-0.76 0.9], p>0.7) so there was no spontaneous remission of symptoms.

Further, significant improvement was observed for secondary outcomes measured at post relative to pre-intervention in trial completers (n=40, Figure 4) for subjective measures of clinician-rated depression (HDRS: d = −1.03, CI [−1.41, −0.65], p<1E-6), self-rated quality of life (MCS12: d = 0.68, CI [0.34, 1.02], p<0.001) and mindfulness (MAAS: d = 0.78, CI [0.42, 1.13], p<0.0001). In addition, objective measures of cognition showed significant improvement in performance efficiency for selective attention (d = 0.51, CI [0.19, 0.84], p<0.001), interference processing (d = 0.53, CI [0.2, 0.85], p<0.01) and working memory (d = 0.66, CI [0.32, 0.99], p<0.001), but not emotion bias (p = 0.3). All significant results were fdr-corrected for multiple comparisons, and there was no effect of iMAP lifestyle domain covariate or participant demographics on any of these measures (p>0.16).

**Figure 4.**
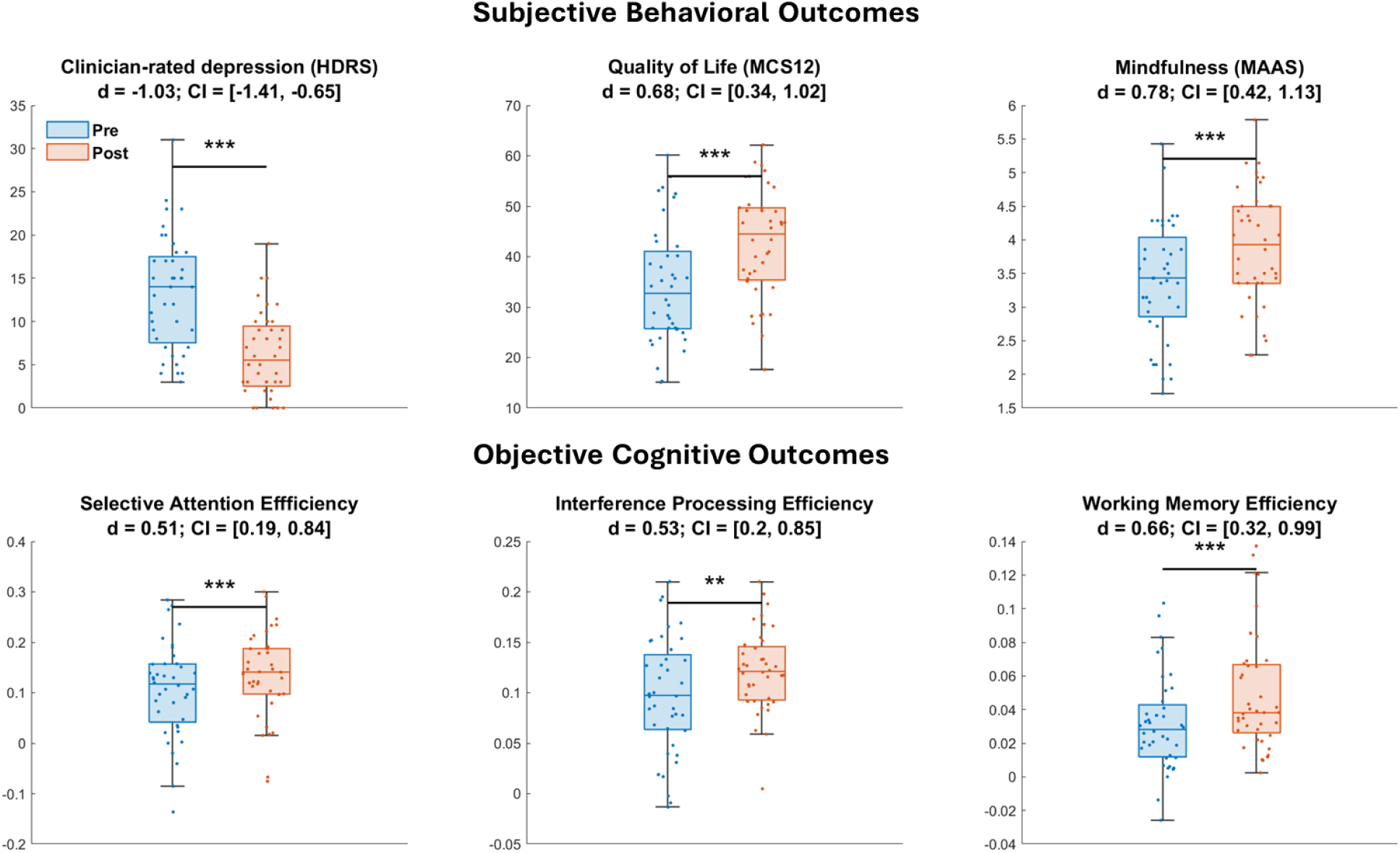
Pre- vs. post-intervention results for secondary outcomes of subjective behavior (top row) and objective cognition (bottom row) in trial completers (n=40) shown as box and swarm plots. Box plots show median with lower and upper quartiles as the box edges, whiskers denote the data range (excluding outliers) and the scatter points show individual participant values. Effect sizes with lower and upper 95% confidence intervals in brackets are shown under each measure label. Stars indicate fdr corrected significance. *: p<0.05; **p<0.01; ***p<0.001

### Relationship between change in depressed mood and iMAP targeted lifestyle domain

Figure 5 shows that iMAP-related alleviation of depressed mood (d = 0.85, CI [0.49 1.2], p<0.0001) was accompanied with improvement in the individually targeted lifestyle domain (d = 0.62, CI [0.28 0.95], p<0.0005) but no significant change in the off-target domains (p>0.3). The evolution of these EMA metrics across phase 2 is shown in Figure 5B. A robust linear regression model assessing the overall relationship between the slope of depressed mood and the slopes of target and off-target lifestyle domain variables across all subjects was significant (F=19.8, p<1E-5) with a significant regression coefficient observed only for the primary target domain slope (β=0.4±0.09, p<0.0005) but not for the off-target slope (p>0.1). For visualization purposes only, the evolution of EMA data binned by GS is also shown (Figure 5C).

**Figure 5.**
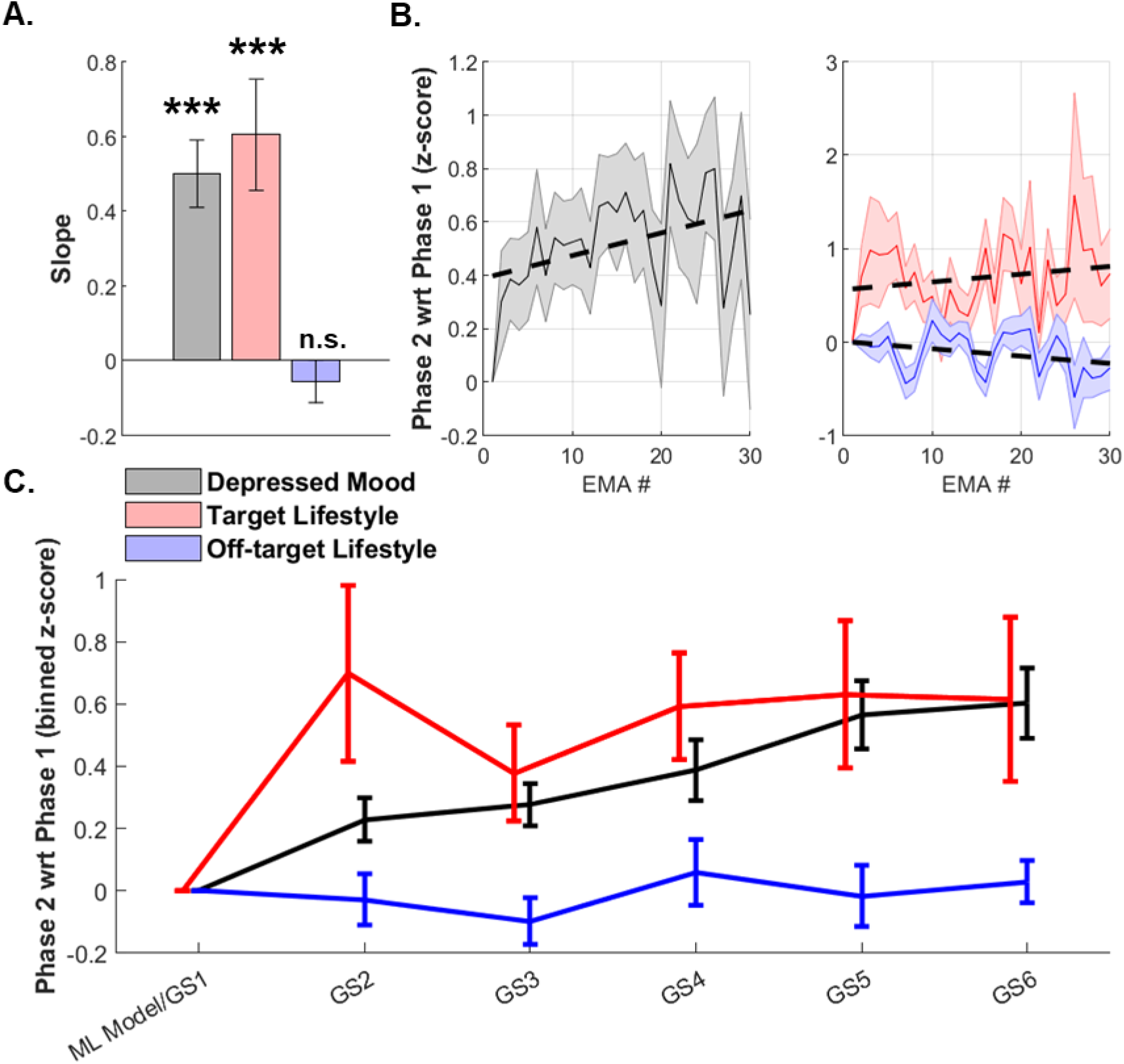
EMA metrics during intervention phase 2. (A) Average slope, i.e., z-scored change in phase 2 relative to phase 1 in depressed mood (grey), target (red), and off-target (blue) lifestyle domains. *** indicates p<0.001; n.s.: non-significant. (B) Evolution of z-scored depressed mood (grey), target (red) and off-target (blue) EMAs across all participants, shaded regions reflect sem. wrt: with respect to (C) Z-scored change in phase 2 relative to phase 1 EMA metrics binned by guided sessions (GS); mean ± sem bars are shown for depressed mood (black), target (red) and off-target (blue) EMAs.

## Discussion

This study introduced the Personalized Mood Augmentation (PerMA) trial as a novel, data-driven N-of-1 ML based behavioral lifestyle intervention approach for mild-to-moderate depression. The trial included a two-week digital monitoring phase acquiring smartphone EMA and smartwatch data, based on which personalized ML was executed and Shapley statistics were applied to reveal lifestyle features rankings that best predict individual mood fluctuations over time. Based on top personalized ML Shapley features, iMAPs were assigned to each participant for a six-week intervention with once-a-week human coach guidance.

Overall the study showed high feasibility with high accuracy best-fit ML models generated for all participants, 100% of participants completed intervention phase 1 (digital monitoring) and 80% of participants completed intervention phase 2 (coach guided iMAPs). Regarding non-completers, main reasons for discontinuation were their busy personal schedules that limited their participation. The 40 PerMA trial completers showed significant alleviation of depressive symptoms (PHQ9, primary outcome) at post-intervention (effect size d = −0.89, CI [−1.25, −0.53]) sustained at 6 weeks (d = −1.06, CI [−1.51, −0.6]) and 12 weeks (d = −0.52, CI [-0.95, −0.09]) follow-ups. Secondary outcomes also showed significant improvement in co-morbid anxiety (GAD7), clinician-rated depression (HDRS), quality of life (MCS12) and mindful attention awareness (MAAS). Additionally, objective cognitive outcomes significantly improved at post-intervention. Here, we discuss several nuances of these results.

The primary PHQ9 outcome and most secondary outcomes showed robust medium-to-large effect sizes for post vs. pre-intervention change. While a control group was absent in this first implementation of the PerMA approach, the effect size comparing post vs. pre-intervention results for phase 1+2 completers (n=40) vs. phase 2 non-completers (n=10) was also large (d = −0.84, CI [−1.54, −0.13]). Intent to treat effect size for PHQ9 scores including all 50 participants was d = - 0.67, CI [= −0.97, −0.36]. The domain of iMAP intervention targeted to either sleep, exercise, diet or social connection did not influence outcomes suggesting that all lifestyle intervention domains were similarly effective when personalized with the caveat that we have limited statistical power to conclude this. It was also interesting to note that social connection was the most implemented iMAP followed by exercise, which may be because these domains had a larger set of actionable variables to model than sleep and diet iMAPs (see Supplementary Methods for actionable variables in each domain). Further, we did not find an influence of demographics of age, gender and ethnicity on trial outcomes.

Notably, while depression can be associated with cognitive deficits ^32–38^, there has been a lack of intervention studies in depression showing robust cognitive benefits. In a meta-analysis of randomized controlled trials, cognitive behavioral therapy (CBT) for depression was confirmed to benefit attention and verbal learning but no other domains of cognition ^68^. There is much evidence that lifestyle intervention and adoption of healthy lifestyles promotes cognitive health and staves off cognitive decline ^69–76^, but again effects of personalized lifestyle optimization for cognition in depression have not been studied. Here, we show that personalized lifestyle intervention significantly improves selective attention, interference processing as well as working memory in individuals with depressive symptoms – all cognitive domains known to be impacted in depression ^77–80^. Moreover, these objective cognitive outcomes complement the subjective behavioral outcomes and confirm broad benefits of personalized lifestyle optimization.

Here, we also confirmed that significant change in the primary targeted lifestyle domain for each individual in intervention phase 2 relative to phase 1, measured using EMA, was significantly linked to improvement in depressed mood. This finding aligns with the strong foundation in the literature that improvements in sleep, diet and exercise are linked to improvements in depression symptom ^81–83^. Changes in off-target lifestyle domains was not significant in phase 2 and not related to improvement in depressed mood emphasizing the specificity of outcomes, i.e., the results were not driven by general lifestyle change across all domains. These results relay the importance of selective personalization of intervention to the lifestyle domain factors that best predict depressed mood in each individual.

The primary limitation of this study is the lack of a randomized control group. Lifestyle targeted behavioral interventions in depression have typically shown small-to-medium effect sizes of ∼0.3-0.5 ^5–17^, while the placebo effect of behavioral interventions are even smaller ∼0.2 ^84,85^. The preliminary effect size of this personalized ML guided trial of 0.8 is similar to the effect size observed for CBT ^86^; yet, an RCT is required for further interpretation of the advantages of personalization over non-personalization. In this context, meta-analytic evidence suggests that psychotherapy personalization is an effective strategy to improve therapeutic outcomes and even small effect size advantages of personalization may have important impacts at a clinical population level ^26^.

Despite its limitations, this trial demonstrates the promise of data-driven personalized lifestyle optimization for improving depressive symptoms, quality of life and cognition. Given the digital health implementation approach, the cost and logistics burden for participants is minimal, and such a lifestyle intervention is further devoid of side effects of pharmacotherapy ^87^. An RCT as the next step will solidify the evidence-base and utility of this data-driven personalized intervention for application at scale.

## Author contributions

Conceptualization: J.M.; methodology: J.N., C.T., D.R. and J.M.; formal analysis: J.N.; investigation: J.N., S.P., S.J., J.K.M., H.A., V.M. D.R. and J.M.; resources: J.M.; data curation: J.N., S.P., S.J., J.K.M.; writing—original draft preparation: J.N.; writing—review and editing: J.N., C.T.T., D.R., and J.M.; visualization: J.N. and J.M.; supervision: J.M.; project administration: J.M.; funding acquisition: J.M.

## Data Availability

All data produced in the present study are available upon reasonable request to the authors

## Acknowledgements

This work was supported by a seed grants from the Hope for Depression Research Foundation (JM). The BrainE software is copyrighted for commercial use (Regents of the University of California Copyright #SD2018-816) and free for research and educational purposes. The machine learning pipeline deployed here is filed s an Invention Disclosure for “Personalized Machine Learning of Depressed Mood using Wearables” (Regents of the University of California Invention Disclosure #SD2021-335).

## Supplementary Materials

### Supplementary Methods

#### S.1 Intervention Phase 1 – Digital monitoring and personalized ML modeling

During this phase, participants wore a smartwatch and completed EMAs of 1-2 minute duration on the BrainE© mobile app’s MindLog module ^1,2^ up to 4X per day for max 60 sessions completed over 2-4 weeks. Participants received daily app notifications sent at 8am, 12pm, 4pm and 8pm until all EMAs were completed.

Ecological Momentary Assessments. EMAs were used to monitor mood ratings and lifestyle factors including sleep, diet, exercise, and social connection as per our prior publications ^1,2^ and as detailed below – Mood Ratings: Participants rated depression and anxiety on 14-point Likert scales shown as green to red color gradient scales. For depression, participants responded to “How happy vs. sad/ depressed are you feeling?” with the “Happy” label anchor next to score of 1 in light green and the “Sad or Depressed” label anchor next to score of 14 in red. For anxiety, participants responded to “How relaxed vs. anxious are you feeling?” with the “Relaxed” label anchor next to score of 1 in light green and the “Anxious” label anchor next to score of 14 in red.

Sleep EMA: Only at their first EMA each day, participants reported their prior night’s sleep time, wake up time, sleep duration, percentage estimate of the time in bed spent asleep (sleep efficiency), and provided a 1–5-star rating of their sleep satisfaction. At EMAs every other time in the day, participants reported nap duration in minutes if they had napped in the past 4 hours.

Diet EMA: At each EMA, participants reported on their recent consumption of sugars, fats, and caffeine in the past 4 hours. To improve compliance, we opted for a simplified version of diet reporting instead of more objective methodologies which can be burdensome ^3,4^. Specifically, within the context of depression, excessive consumption of processed fats and sugars has been related to the severity of symptoms, and intervention to change such diet patterns has shown success ^5–8^. Hence, based on a standard assessment of dietary fats and sugars ^9^, participants responded how many portions (0-12) of each of the following items they had consumed in the past 4 hours: Red meat burger/sandwich, sausage/salami/bacon, whole egg, white bread, pizza, cheese, french fries, chips, butter popcorn, whole milk/milkshake, and fast-food take-out (fats category); cake/cookies, ice-cream, chocolate, candy, pancakes/french toast, jam/honey, soda, juice or other sweetened beverage, and cereal with added sugar (sugars category); and cups of caffeine (coffee/tea/energy drink). Participants also provided a 1-5 star rating of their diet satisfaction.

Exercise EMA: At each EMA, participants reported the amount of exercise (in hours and minutes) they did in the past 4 hours, if any, in these three categories:

a. Strenuous exercise (e.g., running, vigorous sports or bicycling)
b. Moderate exercise (e.g., fast walking, easy bicycling, swimming, dancing)
c. Mild exercise (e.g. yoga, easy walking)

Participants also provided a 1-5 star rating of their exercise satisfaction.

Social Connection EMA: At each EMA, participants reported their social connection in the past 4 hours:

a. Did you chat with family/friends? Yes/No

If yes, how many people close to you did you talk to? 1-10+ how much total time did you spend chatting (in hours and minutes)?

b) Did you attend an organized group in-person or online? (support/sports/exercise/hobby/professional group)

If yes, how long were you engaged (in hours and minutes)?

c) Did you do volunteer work for any organization in-person or online (religious, charitable, political, health-related)?

If yes, how long were you engaged (in hours and minutes)?

Participants also provided 1-5 star rating of their social connection satisfaction.

At end of every first of four EMAs, participants also completed 5 minutes of daily mindful attention to breathing ^10,11^, entered a positive self-reflection and rated their sense of gratitude, and also completed a 30-second attention to breathing assessment at the other three of four daily EMAs; these brief practices were employed to engage mindful awareness and build a foundation to support behavior change in phase 2 ^12–18^.

Daily mindful awareness. At end of every first of four EMAs during the digital monitoring period, participants completed 5 minutes of daily mindful attention to breathing ^10,11^, entered a positive self-reflection and rated their gratitude; these brief practices were employed to engage mindful awareness. In addition, after every three of four EMAs, participants completed a 30-second stress assessment with attention to breathing.

The daily mindful attention to breathing was delivered in a closed-loop game-like format, was performance adaptive and allowed for moment-to-moment performance tracking for quantifying progress and adherence during each session ^19–21^. Specifically, individuals were requested to close their eyes, pay attention to their breathing, and tap the mobile screen after a specific number of breaths. The app monitored the consistency of tap responses. If the user was distracted based on the low consistency of breath monitoring taps, a gentle chime reminded the user to let go of the distraction and revert their attention back to mindful breathing. Initially, at level 1, participants tapped the screen after each breath. If they were able to do this consistently they graduated to tracking 2 breaths at a time, and so on until they were monitoring max 10 breaths at a time at level

10. When the practice ended and participants opened their eyes, they would see a peaceful nature scene slowly unfold as a form of training reward.

For the brief positive self-reflection, participants were prompted with a simple question based on the literature of positive psychology and gratitude ^22^. Question prompts were refreshed every few sessions and included: (1) Who or what made you smile? (2) Who or what are you thankful for?

(3) Note a moment you enjoyed. (4) Note an act of kindness you did or observed. (5) Note a moment you found inspiring. (6) Who or what keeps you going? (7) Note a moment worth celebrating! (8) Everyone has personal strengths. Recognize one of yours. (9) Think of a challenge you faced, small or big, and what you learned from it. (10) Dedicate a note of appreciation to yourself or your loved one(s). Participants completed a brief text response and total time spent on the module as well as active time spent typing out a response was recorded.

Gratitude was rated on a 1-7 Likert scale as a response to the prompt, “Take a moment to indicate how grateful you are feeling.”; participants were given 7 icons graded from rainy weather to sunny weather to choose from.

For the 30-second stress assessment, participants simply tapped the mobile screen after each full breath (inhale plus exhale) ^1^. Recent research shows that consistency of tapping on this basic assay inversely relates to the internally distracted/ruminative state of the individual, which is exacerbated in depression ^23^. Mean breathing response time and consistency data were extracted on this assessment.

Smartwatch data used included heart rate, step attributes (count, speed, distance and calories burned).

Personalized machine learning (ML) Modeling - Data Ingestion, Feature Extraction, Personalized ML Pipeline. Data from both smartphone and smartwatch sources were carefully aligned. For this all independent data variables were either aggregated or extrapolated based on their sampling frequencies to match the sampling frequency of the dependent variable (DV), i.e., depressed mood EMA ratings. All features extracted from the EMA and smartwatch data are enumerated below; the personalized ML pipeline was adopted from our previous research ^1,2^.

EMA data -

1. Time of the day when a particular dependent variable (DV, i.e., depressed mood rating) was taken: (0:00, 10:00), (10:00, 14:00), (14:00, 18:00), (18:00, 23:59)
2. Anxiety ratings completed at each time point when a DV rating was obtained.
3. Sleep time, wake up time, sleep duration, percent estimate of time in bed spent asleep, and 1– 5-star rating on sleep satisfaction the previous night.
4. Exercise duration for each intensity type and total satisfaction in the 24 h period prior to each DV rating.
5. Total amount of fats, sugars, caffeine, and diet satisfaction in the last 24 h of each DV rating.
6. Number of people and total time spent chatting, total time in an organized group, total time spent volunteering, and total satisfaction in the 24 h period prior to each DV rating.

Mindful awareness module data –

7) Attention to breathing mean time and consistency obtained at each DV rating.
8) Gratitude rating completed every fourth DV rating.
9) Total and active response time in the positive self-reflection completed every fourth DV rating. Smartwatch data –

10) Heart rate taken as the mean value within a ±30 min window around the time of each DV rating.
11) Cumulative step features taken as the mean values from the past 12 h of each DV rating for each step feature separately, i.e., count, speed, distance and calories burned.
12) Cumulative exercise features taken as the mean values from the past 12 h of each DV rating calculated for each feature separately, i.e., duration and calories burned.

The modeling pipeline is shown in Supplementary Figure 1 below. The first step prior to ML modeling involved data ingestion, feature extraction and data preprocessing. For this the above data features were extracted from each participant’s EMA, mindful awareness module and smartwatch data.

**Supplemental Figure 1.**
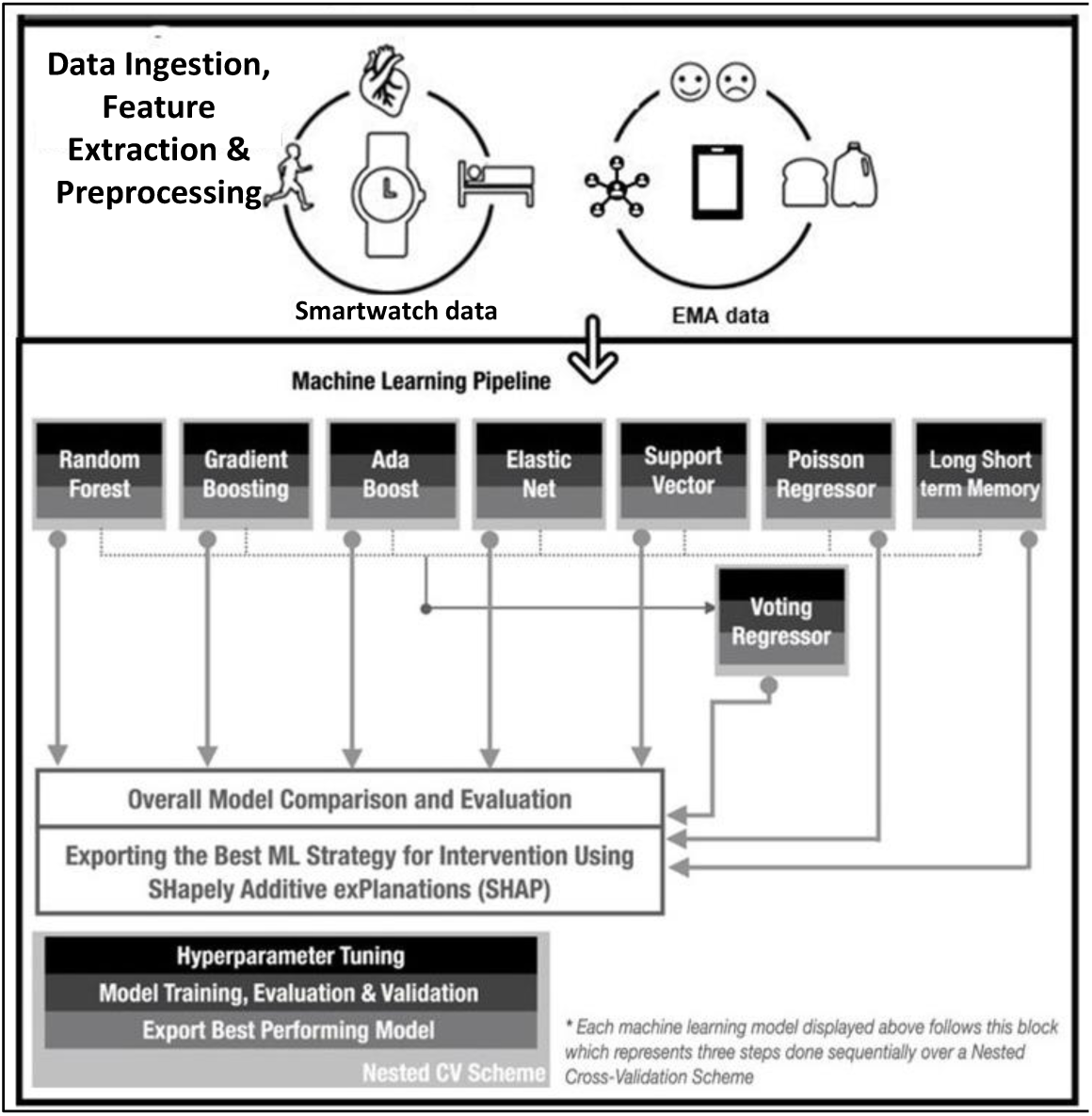
Flow of the personalized Machine Learning (ML) modeling pipeline ^1,2^.

Features which participants had no responses for through the entire study were considered missing and dropped for that participant. All features were calculated and stored separately for each subject for a max of 34 possible features per participant. Data were also inspected using both automated and manual approaches for unusable and missing variables, as well as variables with zero variance that were dropped for that participant. We did not implement any additional feature selection such as principal component analysis, which would dissociate variables from their physical attributes, to preserve model interpretability. Manual inspection of EMA data was also done in cases where data was not delimited properly, and to correct any data values that were saved in the wrong units (i.e., using hour fractions instead of hour and minute representations for durations). Other manual inspection of raw data was only used to verify meta data file names, variable names, and data format differences that occurred from different mobile operating systems and smartwatch versions.

Data preprocessing took participant data matrices from the prior step for purposes of imputation, standardization, and regularization. The preprocessing ensured to not alter the overall distribution of the data at the level of each participant. Iterative imputation was used for any missing data. For personalized models, removing missing data can create unaccountable bias and lead to low accuracy on test data. Moreover, filling missing values with fixed values, mean, mode, or median can also cause problems; when filled in place of missing data, these values can alter the original multivariate distribution, which may hinder the model from generalizing actual patterns in the training dataset. Thus, for missing data, we used iterative imputation, a regression-based multivariate imputation scheme ^24^. This scheme models each feature with missing values as a function of other features and uses that estimate for imputation. It does so in an iterative round-robin fashion: at each step, a feature column is designated as output Y, and the other feature columns are treated as inputs X. A regressor is fit on (X, y) for known Y. Then, the regressor is used to predict the missing values of y, executed for each feature in an iterative fashion. This iterative imputer is referred to as Multivariate Imputations via Chained Equations (MICE) ^1,2^.

To achieve preprocessing efficiency over computationally heavy ML processes, a preprocessing pipeline object was used. Using such an object has various advantages, including but not limited to encapsulating the preprocessing steps together, and avoiding leaking statistics from the test data into the trained model in cross-validation (CV) by ensuring that the same samples are used to train the transformers and predictors, and improving run time during parallel processing. For this study, the following preprocessing pipeline strategy was devised: (a) continuous and discrete variables were processed independently, (b) discrete variables were imputed using a "most frequent class imputer", which is essentially filling missing values with the class with highest frequency, (c) the continuous variables were imputed using the MICE method described above, (d) all discrete variables were regularized using an ordinal encoder, which results in a single column of integers (0 to n-categories - 1) per feature, and finally (e) all continuous variables were regularized using a maximum absolute scaler, which scales and translates each feature individually with the maximum absolute value in the training set such that it does not shift or center the data, thereby, not destroying any sparsity. The data was then ready to be deployed in the ML analysis pipeline.

ML Pipeline. A primary step to achieving robust ML models is ensuring independence between training and test, and providing transparency on the models that are evaluated. The personalized ML pipeline included hyperparameter tuning, model training, evaluation, and model selection. On the one hand, ensuring independence between data, which is used for hyperparameter tuning, training and testing makes the model less prone to overfitting, and prevents the introduction of bias into the model. However, ensuring independence between training and test datasets is a particular challenge for the N-of-1 modeling approach. A traditional k-fold cross-validation (CV) scheme cannot be used in this case as the model performance would then be highly dependent on the small number of examples set aside for testing. Thus, to tackle this technical challenge and achieving a model practically free from bias and immune to overfitting, a nested CV scheme was used ^25–29^. Specifically, we used a repeated 4-fold CV scheme with ten repeats as the inner CV strategy and a simple 4-fold CV scheme as the outer CV strategy for the overall nested CV scheme. We then modeled individual depression ratings based on the various EMA and smartwatch lifestyle predictors, employing supervised ML regression models hyperparameter tuned and trained over the nested CV scheme. The main steps of the pipeline include comparison of multiple ML strategies for each subject including Random Forest, Gradient Boost, Adaptive (Ada) Boost, Elastic Net, Support Vector, Poisson Regressor and Long Short-term Memory. The Voting Regressor employed the best model from all the other ML strategies besides LSTM. After hyperparameter tuning and training over all ML models, results were evaluated for each model, and each subject over the regression metrics of mean absolute percentage error (MAPE). We used MAPE as the performance metric to choose the best model (with lowest error) for each ML strategy. The best model for each strategy was then fed in the voting regressor. The best model from this strategy was calculated in the same manner as the other strategies. We then compared the outcome of the best performing models from each ML strategy and calculated the overall best model with the least overall MAPE. Thus, each study participant would have their own personalized model predicting their depressed mood fluctuations over time.

For each person’s best-fit personalized ML model we utilized Shapley statistics, specifically SHapley Additive exPlanations (SHAP), a widely used game theory based algorithm, to explain feature importance ^30^. Shapley values indicate the relative importance and directionality of each feature as it predicts the depressed mood DV ^1,2^. Features with large absolute Shapley values are essential, hence, we rank-sorted these feature values and the top 10 (that accounted for majority of the variance attributed to the DV by the model) were inspected by the coaches to determine individual Mood Augmentation Plans (iMAPs). Examples of Shapley plots that map to the 4 iMAP domains are shown in the main text Figure 2. The personalized ML model results, thereby, revealed the specific lifestyle attributes that are most impactful in predicting each individual’s depressed mood, and the top-ranking predictors could then be used to guide each participant’s iMAP.

#### S.2 Intervention Phase 2 – coach guided iMAPs

Behavioral coaches reviewed the personalized ML Shapley results to determine the top-priority lifestyle intervention domain (i.e., sleep/ exercise/ diet/ social connection) for each iMAP. In case features from more than one domain were present within the top 10 predictors of depressed mood, coaches also based their decision on number of domain features present as top predictors, whether these predictors independently showed high correlations with depressed mood, whether participants reported low satisfaction with the chosen lifestyle domain, and whether it was feasible for participants to initiate intervention in the chosen domain (e.g., fixed sleep schedules due to work may constrain a person’s ability from engaging in a sleep-focused plan, in which case the second most mood-predictive domain would be the focus of intervention). Description of each 6-week iMAP in either sleep/ exercise/ diet/ social connection domain is listed below and grounded in evidence-based intervention protocols in these lifestyle domains in the context of depression ^6,7,22,31–35^.

Sleep iMAP. This plan was chosen if low sleep efficiency and low sleep satisfaction (actionable features in the sleep domain) were top predictors of an individual’s depressed mood. The sleep iMAP is based on cognitive behavioral therapy for insomnia (CBT-I) that has shown medium effect size improvements (d=0.5, confidence interval = 0.3-0.8) for depression treatment ^36,37^. Guided session (GS) 1 focuses on review of the personalized ML model to show top-ranking sleep predictors for depressed mood. The health coach then introduces the concepts of CBT-I including sleep hygiene and stimulus control, waking up at a regular time and avoiding/reducing naps. Participants keep daily electronic sleep logs that are then reviewed each week by the health coach. In GS2, the concept of sleep efficiency is introduced and the coach reviews whether the participant is keeping up with sleep hygiene guidelines. In GS3, daily sleep log review continues and if high sleep efficiency is achieved, then time in bed is incrementally increased. The participant also completes a SMART (Specific and small, Measurable, Action oriented, Realistic, Time stamped) goals assessment for sleep hygiene. GS4 focuses on thoughts around sleep and encourages scheduling worry time during the day (not in bed). It also introduces practicing cognitive restructuring around negative sleep thoughts (Example: “If I can’t get a good night’s sleep my day tomorrow will be shot”; Restructured: “Even if I don’t sleep well tonight, I can still get up in the morning and do things. And the more active I am tomorrow, the easier it will be to fall asleep tomorrow night.”). GS5 is dedicated to continued review of the daily sleep log, any difficulty with sleep hygiene and restructuring sleep thoughts, and titrating time in bed based on sleep efficiency over the past week. GS6 continues to review prior sleep log progress and focuses on relapse prevention by assigning an action plan to address insomnia in the future.

Exercise iMAP: This plan was chosen if exercise features were top predictors of an individual’s depressed mood. Actionable feature variables in the exercise domain included cumulative step distance, cumulative step speed, cumulative step calories, cumulative step rate, heart rate, exercise satisfaction, exercise calories, exercise duration, past day mild/moderate/strenuous exercise. Like the sleep intervention, exercise has also shown moderate effect size improvements in depression per systematic review and meta-analysis of exercise randomized control trials (RCTs) ^38^. In this iMAP, participants first review their personalized ML model for daily exercise features as top-ranking predictors of depressed mood. In GS1, the coach discusses what a consistent exercise training plan would look like for the participant. The coach uses a motivational interviewing strategy to discuss where participants are at in their change process (pre-contemplative, contemplative, ready for action) and identify specific obstacles that have prevented change in the past. As 150 minutes/week of exercise are recommended for general health ^39^, the program’s goal are to help participants to progressively build up to that level. At GS1, the participant is also requested to completed a SMART exercise solutions assignment in which the participant identifies challenges to regular exercise and feasible solutions to these. In GS2-6, the coach reviews the participant’s exercise EMA and smartwatch logs as well as SMART exercise solutions. The coach encourages and amplifies positive progress made and discusses any revisions to the SMART exercise solutions, if necessary. The participant is encouraged to increase exercise target goals by 20% each week until participant reaches about 150 mins/week of consistent activity. This activity is also objectively monitored with smartwatch data.

Diet iMAP: This plan was chosen based on diet variables, including diet satisfaction, past day fats, and past day sugars (actionable features in the diet domain) as top predictors of depressed mood. This plan is based on the modified Mediterranean diet that has shown evidence for efficacy in depression alleviation ^40,41^. In GS1, the coach reviews the participant’s personalized ML dietary predictors of depressed mood and general patterns of poor eating and diet dissatisfaction. Similar to the exercise plan, motivational interviewing is used to identify barriers to diet change, and that the goal is to re-align eating as close as possible to the target healthy mood diet. Contents of the healthy mood diet are introduced including servings of whole grains, fruits and vegetables, nuts, legumes, fish, eggs and olive oil. The diet recommends to reduce dairy, red meat and poultry servings and avoid sweets, processed cereal, chips, pastries, fried food, fatty meat, dairy, desserts, sugary drinks, condiments and alcohol. Weekly eating tips are provided along with convenient meal and snack ideas and example meal plans. A healthy mood diet grocery shopping list is also provided. Participants are encouraged to complete a healthy mood diet log that is reviewed weekly. In session 2-6, the coach reviews the participant’s healthy mood diet log and the participant completes a SMART eating solutions assignment. The coach also discusses how tangible change can be progressively achieved, for instance, focusing on changing one meal a week (i.e. breakfast/lunch/snack/dinner) instead of all meals at once. The final session focuses on maintaining the consistent progress made with the healthy mood dietary changes.

Social Connection iMAP: This plan was chosen if number of people and total time spent chatting (connect chat people and chat time features), total time in an organized group (connect group time), total time spent volunteering (connect volunteer time), connection satisfaction, gratitude and active reflection time (all actionable features in the social domain) appeared as top predictors of depressed mood. This iMAP is based on the behavioral intervention for positive amplification of mood that has shown improvements in positive affect and social connectedness ^42,43^. The intervention comprises 3 core elements: 1) increasing exposure and responsiveness to positive events; 2) practicing gratitude; and 3) engaging in kind/ generous acts toward others. In GS1, the coach reviews the participant’s personalized ML and specific social connection predictors of depressed mood, and introduces the three core elements of the positive amplification of mood plan. The participant is given a goal setting assignment to note down what’s important to them in life and instructed to complete a positive event tracker at least once per day. The positive event tracker asks to describe the event and associated emotions, physical reactions, intensity, duration and individual response. GS2 continues to emphasize increasing exposure and responsiveness to positive events as well as completing a weekly gratitude reflection assignment describing up to 5 things the participant is grateful for. GS3 continues prior activities and adds on acts of kindness for others. During one day in GS3, the participant is requested to perform up to five acts of kindness – all in one day. These acts do not need to be for the same person, and the person(s) may or may not be aware of the act. Once completed, the participant reports on the acts of kindness – what they did and how they felt before, during and after the completed act. GS4 also builds on prior weeks and focuses participants to schedule pleasurable, engaging, and meaningful activities. The participant is encouraged to complete a pleasurable activity alone as well as with others. The participant must also complete an engaging activity in which s/he tends to lose awareness of time or sense of self such as completing a task that challenges skills (e.g., playing a new piece of music; walk or run; engaging in an important yet challenging social interaction or event). And finally, the participant must complete a meaningful activity such as completing a challenging task that one is avoiding, or helping others with a challenging task. The participant logs the pleasurable activities done alone and with others, and also logs the weeks engaging activity and meaningful activity – the emotions and reactions these activities generated and what the individual noted. GS5 continues on building prior progress and introduces creation of positive self-perpetuating interpersonal cycles. In this activity, the participant identifies people within their social network that they would like to further develop or strengthen their relationship with. These could be new relationships (e.g., acquaintances) or existing relationships with friends or family. The participant is then encouraged to use a combination of strategies from previous sessions (increasing responsiveness to positive events, gratitude, and engaging in prosocial acts) to strengthen their relationships. The participant is encouraged to develop a positive activities plan for the future that incorporates activities they liked most, integrating variety, identifying the optimal duration and timing of activities, and involving others for support. Finally, GS6 focuses on relapse prevention. Participants review what they learned in the coaching, how they will maintain and build upon their gains, discuss potential obstacles and establish a plan for addressing them.

During this phase, each participant met once-a-week with their coach for a ∼20 minutes GS video call to discuss progress and any impediments to behavior change. Participants also completed once-a-day EMAs (same as those in phase 1) over the 6 weeks to track progress.

#### S.3 Automated iMAP lifestyle domain assignment

While artificial intelligence (AI) chatbot based therapy is gaining traction ^44–46^, the depth of understanding and empathy provided by a human coach are still unmatched by chatbots ^47–49^, hence, we decided to integrate human coaches that had remote weekly video check-ins with each patient. However, advancements in AI can be leveraged in another manner. In order to further streamline and automate the iMAP assignment, we generated a decision algorithm (DA) as well as an LLM prompt that took into account all factors of the personalized ML results, which were considered by the coaches to recommend specific iMAPs. The DA followed the same criteria as human assignment, see equation 1 below where a higher domain score meant a more suitable iMAP target domain (i.e., sleep/ exercise/ diet/ social connection) -

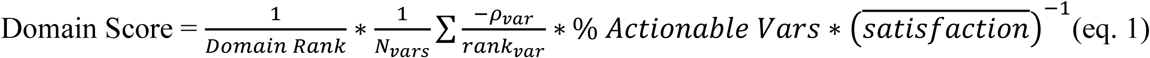

This equation has 4 major parts:

1. 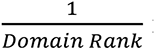: Here, domain rank is 1-4 based on the order they appear in the top 10 SHAP feature list. Lower numeric value means better result, so the operation is inversed.
2. 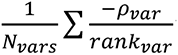: The summation takes into account the independent correlation between each feature variable and depression and scales it by the relative SHAP rank. Lower ranks should be less weighted hence the division, and large negative correlations are desired (i.e., higher values of the feature variable associated with lower depressed mood), hence the negative weight.
3. % *Actionable Variables*: ratio of actionable variables within a domain present in the top 10 SHAP feature list and the total number of actionable variables in the domain.
4. 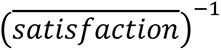: We wanted to prioritize domains with low satisfaction so an inverse relationship was used.

The basic domain score equation did not assign any specific weights to the 4 parts above. We also built a further fine-tuned model by iterating the weights of the parts in the above equation based on the target domain preferences made by the coaches across all study participants. This fine-tuned model is shown in equation 2 below. This equation aimed to maximize the domain score by setting all feature correlations in a domain to the max observed correlation (−𝜌_𝑚𝑎𝑥_) and assigned a weight of 0.9 to this part of the equation. It further added the inverse of lifestyle domain satisfaction to the equation but with a lower weight of 0.1. This fine-tuning was simply done empirically to maximize the percent match in DA-based lifestyle domain assignment with that of the human coach. The domain rank list for the fine-tuned model for all 40 participants is shown in Supplementary Table 1 below.

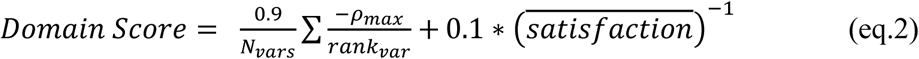

While a decision equation outputs a domain rank list, the additional benefit of an LLM is that it can output logical explanations that may further assist the human coach. However, the LLM will always be a “black box” model even if the prompt provided is transparent and structured. When the naïve decision model aligns with the LLM in the domain ranking order, we harness the LLM’s nuanced, context-rich insights while confirming it adheres to the essential decision equation principles. Thus, the prompt given to the Gemini LLM was also based off equation 1 above and the decision making process of the human coach, and is detailed below. Relevant personalized ML model results data was converted into a JSON file and included with the prompt. Notably, maintaining participant privacy and security, the prompt did not include any personal health information (PHI) for any participant –

You are a behavioral health coach that is helping individual patients pick a domain to work on based on their lifestyle data. The data is split into two categories, when the individual experiences high and low depression.

Data Structure:

- ’rank’: A numerical identifier for each variable indicating importance. 1 means most important

- ’level_0’: Represents the variable name and units.

- ’level_1’: Subcategory within each ’level_0’, including "MEAN", "COUNT", "STD", and "CI95".

- ’MEAN’: Average value.

- ’CORR’: Spearmans correlation of variable to depression. This is the same for both low and high groups

- ’STD’: Standard deviation.

- ’CI95’: 95% confidence interval.

- Relationships:

- Each ’level_0’ value has multiple ’level_1’ values.

- Each ’level_1’ value has corresponding mean, count, std, or CI95 values for the low and high groups.

Domains and relevant variables: The variables belong to one of four possible domains: sleep, exercise, diet, positivity. Ignore all variables that are not listed here.

- Sleep: ’Sleep_percent’, ’Sleep_satisfaction’

- Exercise: ’cumm_step_distance’, ’cumm_step_speed’, ’cumm_step_calorie’, ’cumm_step_count’, ’heart_rate’, ’Exercise_satisfaction’, ’exercise_calorie’, ’exercise_duration’, ’past_day_exercise_moderate’, ’past_day_exercise_mild’, ’past_day_exercise_strenuous’

- Diet: ’Diet_satisfaction’, ’past_day_fats’, ’past_day_sugars’

- Positivity: ’Connect_chatpeople’, ’Connect_chattime’, ’Connect_grouptime’, ’Connect_volunteertime’, ’Connect_satisfaction’, ’Gratitude’, ’Reflect_activetime’

IMPORTANT: You must only use the variables specified above when providing justification. Do not include any variables that are not listed.

Your task is to:

Rank the domains from 1-4. This ranking should be weighted based on the ranking of the highest variable, proportion of relevant variables relative to all variables possible, and strong negative correlation to depression.

Analyze the provided data to rank the intervention domains for behavioral change and suggest specific, actionable recommendations. Recommendations should prioritize positive lifestyle modifications. Rankings for intervention domains for behavioral change should be determined based on number of, rank, and correlation strength of impactful variables that show up.

1. Parameters:

– Do not extrapolate any information that is not given
– If a domain has no relevant variables, it should automatically be ranked last
2. Prioritization:

– Higher ranked variables should be weighted more
– Any domain with variables that have a positive CORR value should incur a penalty
– Variables with stronger negative correlations should be given priority. <-0.4 is considered strong, between [-0.4, −0.2] is considered medium, and between [-0.2 and 0] is considered weak
– Give priority to domains where multiple variables are significant.
3. Suggestions:

– Align with the ranked importance of variables.
– Promote healthy lifestyle changes.
– Ensure recommendations are evidence-based and feasible for implementation.
4. Justification:

– Provide an analysis for each variable based on rank, correlation strength, penalties and how it affects the overall rankings.

Requirements:

- rank the Four Intervention Domains in order by effectiveness of domain: sleep, diet, exercise, or positivity.
- Specific, actionable recommendations that are not drastically different from the current lifestyle.
- Justification referencing data points and variables.

Keep your responses condensed. Provide just the ranking.

#### S.4 Cognitive Assessments

At pre and post-intervention, participants engaged in ∼30-minutes of cognitive assessments on the BrainE© platform. Participants engaged in four assessments of (1) selective attention, (2) interference processing, (3) working memory, and (4) emotion bias, and had the opportunity to take self-paced breaks between tasks to minimize fatigue. Tasks were run in the same order for all participants.

**Supplementary figure 2** shows the stimulus sequence in each task. All four cognitive tasks had a standard trial structure of 500 ms central fixation “+” cue followed by a task-specific stimulus presented for task-specific duration and with a task-specific response window. This response window in each task was adaptive with a 3up-1down staircase scheme that maintains accuracy at ∼80% and engages the user by avoiding ceiling performance ^50,51^. An adaptive scheme also reduces practice effects that affect repeat assessment sessions. Further details of the adaptive scheme in each task are provided below.

**Supplementary Figure 2.**
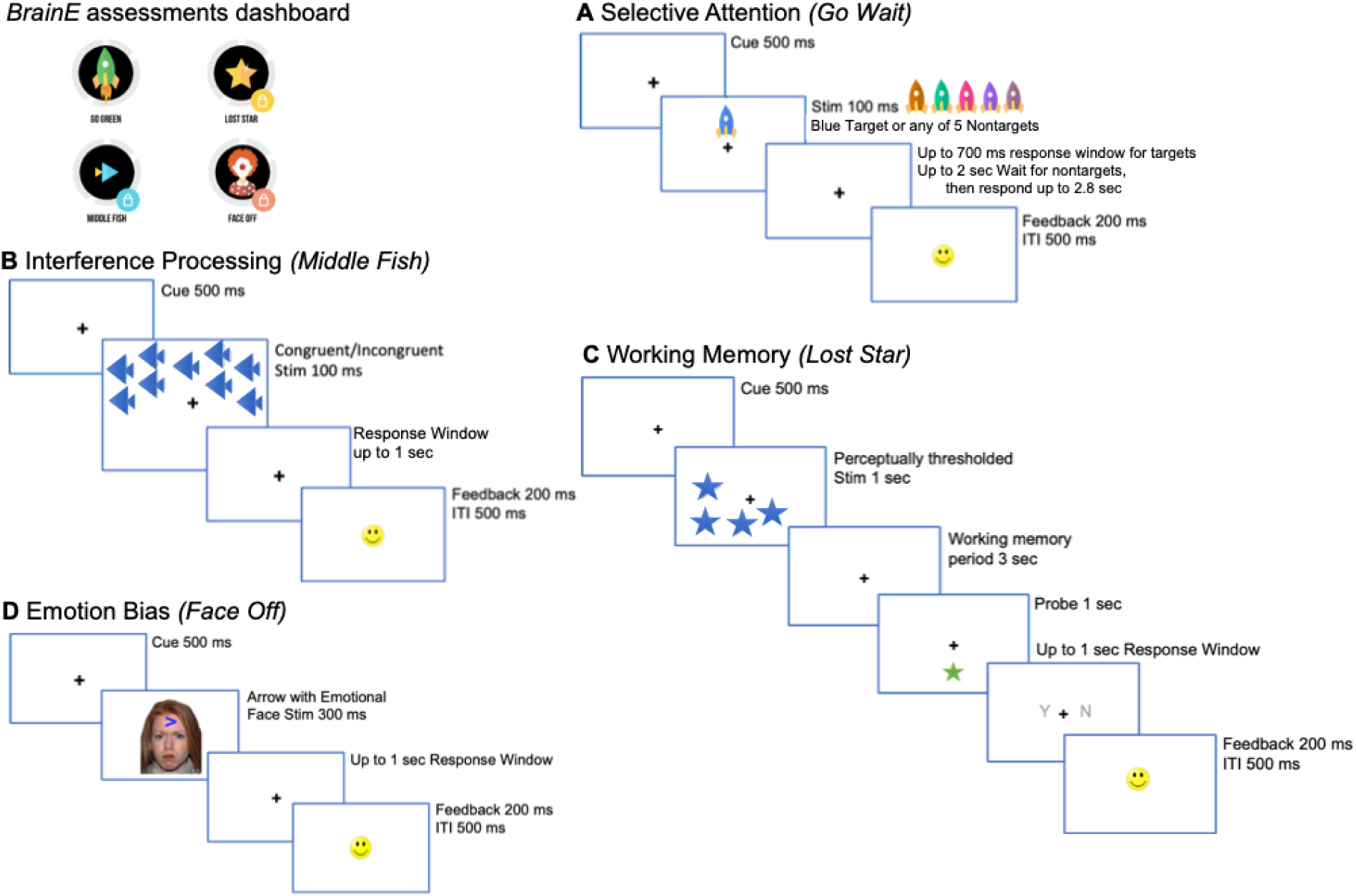
Cognitive assessments delivered on the BrainE platform. At top left, the BrainE assessment dashboard is shown. (A) Selective attention was measured on the ‘Go’ trials of the Go Wait task that required rapid and accurate responses to blue rocket targets. All other color rockets were non- targets on which participants waited to respond for up to 2 sec. (B) In the Flanker interference processing Middle Fish task, flanking fish may either face the same direction as the middle fish on congruent trials as shown, or the opposite direction on incongruent trials; participants were instructed to respond to the direction of the middle fish. (C) The visuo-spatial working memory task, Lost Star, was presented with perceptually thresholded stimuli and participants responded after a working memory period whether a probe star was positioned in one of the same locations as the prior test star stimuli. (D) The emotion bias task, Face Off presented neutral, happy, sad, or angry faces superimposed on an arrow, whose direction was discriminated by participants.

Stimuli in each cognitive task were presented in a shuffled order across trials. Response in every task trial was followed by standard response feedback for accuracy as a smiley or sad face emoticon, presented 200 ms post-response for 200 ms duration, followed by a 500 ms inter-trial interval (ITI). ITI jitter within tasks was not applied to keep the task administration rapid. At the end of each task block, participants received a percent block accuracy score with a series of happy face emoticons (up to 10) to promote engagement.

1. Selective Attention: Participants accessed a game-like task, Go Wait modeled after the standard test of variables of attention ^52^. On each task trial, colored rockets were presented in the upper or lower central visual field. Participants were instructed to respond as rapidly as possible to blue colored rocket targets and wait to respond for 2 sec to distracting rockets of five other iso-luminant colors (shades of brown, teal, pink, purple). Iso-luminant colors were ensured using luminosity measurements in Photoshop. Luminance values for iso-luminant stimuli were 128, estimated as 0.30*R+0.59*G+0.11*B (where RGB are the Red/Green/Blue values of the chosen stimulus color). Post-fixation cue, a target/non-target stimulus appeared for 100 ms duration. For the blue rocket targets, the initial response window was set at 700 ms that adapted on each trial in a 3up-1down scheme, i.e., the response window reduced −33 ms after correct trials and increased +100 ms after incorrect trials. One happy face emoticon followed correct trials. If response was very rapid within 100-400 ms, then two happy face emoticons were presented for feedback to reinforce fast and accurate responding ^53^. For nontarget rockets, response times were not adaptive; participants waited for 2 sec at which time the fixation cue flashed briefly for 100 ms and then participants responded. Across two blocks, target and non-target trials were shuffled with 50% probability for a total of 180 trials. Response efficiency, i.e., the product of accuracy and speed was taken as the main selective attention outcome measure ^54,55^; here, accuracy was measured as the signal detection sensitivity, d’, computed as z(Hits)-z(False Alarms) ^56^ and task speed was calculated as log(1/RT), where RT is the average response time across attended trials in milliseconds.
2. Interference Processing: Participants accessed the game-like task, Middle Fish, an adaptation of the Flanker assessment ^57–59^. Post-fixation on each trial, participants viewed an array of fish presented in the upper or lower central visual field for 100 ms. On each trial, participants had up to a 1 sec response window to detect the direction of the middle fish in the set (left or right) while ignoring the flanking distractor fish that were either congruent or incongruent to the middle fish, i.e., faced the same or opposite direction to the middle fish. Response windows were adapted on congruent trials in a 3up-1down scheme (-33 ms after correct trials and +100 ms after incorrect trials) and incongruent trial response windows matched that of the previous congruent trial. The fish flanking the middle fish interfere with the discrimination of the direction of the middle fish (left/right), hence, the task assesses interference processing. Task trials were shuffled with congruent/incongruent distractors in 1:1 ratio in 96 trials over two blocks. As in the selective attention task, response efficiency, i.e., the product of accuracy (i.e., signal detection sensitivity: d’) and speed was taken as the main outcome measure.
3. Working Memory: Participants accessed a game-like task, Lost Star, which was based on the visuo-spatial Sternberg task ^60^. Post-fixation cue on each trial, participants viewed a spatially distributed test array of objects (i.e., a set of blue stars) for 1 sec. Participants were required to maintain the locations of these stars for a 3 sec delay period utilizing their working memory. A probe object (a single green star of 1 sec duration) was then presented in either the same spot as one of the original test stars, or in a different spot than any of the original test stars. The participant was instructed to respond whether the probe star had the same or different location as one of the test stars. 50% of task trials had the same probe star location as one of the test stars while 50% had different location, and presented in shuffled order. For each participant, we implemented this task at the threshold perceptual span, which was defined by the number of test star objects that the individual could correctly encode without any working memory delay. For this, a brief perceptual thresholding period preceded the main working memory task, allowing for equivalent perceptual load to be investigated across participants ^58^. During thresholding, the set size of test stars increased progressively from 1-8 stars based on accurate performance where 100% accuracy led to an increment in set size; <100% performance led to one 4-trial repeat of the same set size and any further inaccurate performance aborted the thresholding phase. The final set size at which 100% accuracy was obtained was designated as the individual’s perceptual threshold. Post-thresholding, the working memory task presented 48 trials over two blocks. Unlike the two previous tasks, this task was not speeded but instead adapted the working memory period in a 3up-1down scheme; if correct, the working memory period increased by +0.9 sec or if incorrect, it decreased by −0.3 sec. As in the above two tasks, response efficiency was taken as the main outcome measure but also weighted by each person’s item span, i.e., the product of item span, accuracy (i.e., signal detection sensitivity: d’) and speed was taken as the main outcome measure.
4. Emotion Bias: Participants accessed the game-like assessment, Face Off, adapted from studies of attentional bias in emotional contexts ^61,62^ The task integrated a standardized set of culturally diverse faces from the NimStim database ^63^. We used an equivalent number of male and female faces, each face with four sets of emotions: neutral, positive (happy), negative (sad) or threatening (angry), presented on equivalent number of trials in each task block. Post-fixation cue on each trial, participants viewed an emotional face with a superimposed arrow of 300 ms duration. The arrow occurred in either the upper or lower central visual field on equal number of trials. Participants responded to the direction of the arrow (left/right) within an ensuing 1 sec response window. For neutral emotion trials, this response window adapted in a 3up-1down scheme (-33 ms after correct trials and +100 ms after incorrect trials). All other emotion trials followed the same response window as their previous neutral emotion trial. This task evaluates emotion bias (or interference) as the emotional faces interfere with the discrimination of the direction (left/right) of the arrow on which they are superimposed. Participants completed 144 trials presented over three equipartitioned blocks. Again, response efficiency, i.e., the product of overall accuracy and speed was monitored as the main outcome measure.

#### S.5 Supplemental Data Analyses

Majority of our participants completed survey requirements on time, however we had a small number of missing data points for PHQ9 and GAD7 which needed to be accounted for in our analysis. One participant was missing screening data, 2 were missing GS3, and 8 were missing GS5 timepoints for PHQ9, resulting in 4.6% missing. One participant was missing pre data, 2 were missing GS3, and 7 were missing GS5 for GAD7, resulting in 5% missing. Given the limited number of missing data points and their sequential nature, imputation was done independently for each participant with linear interpolation. As these survey data distributions were non-normal, the non-parametric Friedman’s repeated measures test was used to investigate significant change from screening to post-intervention. Sphericity was checked with Mauchly’s test and p values were corrected per the Greenhouse-Geisser method. Follow-ups at 6 and 12 week had more missing data at 30% and 45% missing for both PHQ9 and GAD7. As such, they were not included in the Friedman’s analysis nor imputed. Instead, these time points were assessed with the signed rank test against the pre-intervention time point without imputation.

To assess pre vs. post-intervention differences in secondary outcomes, i.e., clinician-rated depression (HDRS), quality of life (MCS12), and mindfulness (MAAS), normality of each variable was first verified with the Anderson Darling test. Non-normal variables were tested for post vs. pre differences using the signed rank test, else a paired t-test was used.

In an effort to harmonize our primary and secondary outcomes, we conducted a robust linear regression model to investigate the relationship between the post vs. pre-intervention change in the primary PHQ9 outcome and all secondary outcomes controlling for demographics of age, gender and ethnicity. Robust regression was used to minimize any outlier influence ^64^. PHQ9 change from pre-intervention to follow-ups was also similarly investigated while controlling for demographics.

Additionally, we wanted to investigate whether any baseline metrics could serve as indicators of remission. We assessed whether any demographics (age, gender, ethnicity) or secondary behavioral (GAD7, HDRS, MCS12, MAAS) or cognitive outcomes at baseline (i.e., at pre-intervention) related to remission, i.e., post-intervention PHQ9 score <5 reflecting healthy, non-depressed status. For this, we used one-way ANOVA with remitted vs. non-remitted participants as the between-subjects factor for all continuous variables, except categorical variables of gender and ethnicity that were tested using 𝜒 ^2^ test. All p-values were fdr-corrected for multiple comparisons across demographic and baseline outcomes. We further verified that measures were still significant when accounting for baseline depression symptoms in a generalized linear regression model.

Representative Domain Variables for EMA Analysis (During Phase 2). In order to complete this analysis, we needed representative variables to objectively measure behavioral change in each domain. We only considered EMA, and not smartwatch data for this analysis as some domains, specifically diet and social connection, had no corresponding smartwatch metrics. Specific intervention-relevant EMA targets for this analysis were as below (higher values are better) - Sleep: percentage of time spent asleep at night compared to phase 1

Exercise: total time (hrs) engaged in mild, moderate, and strenuous exercise compared to phase 1 Diet: reduction in fats and sugars (portions) consumed compared to phase 1

Social Connection: total time (hrs) chatting, in a group, and volunteering compared to phase 1

From the above, each participant had one primary target metric corresponding to their assigned iMAP lifestyle domain. Lifestyle domains not assigned for intervention to the participant are then referred to as their off-target domains. Up to 30 EMAs were obtained during intervention phase 2 corresponding to 6 weeks of intervention with EMA logs made 5 days per week. To facilitate comparison, all depressed mood EMA as well as all domain metrics in each participant were z-score standardized based on their phase 1 EMA data. Off-target domain metrics were averaged together for a combined representation. Any missing EMAs of 30 were replaced with NaNs. Average z score and shaded error bars representing standard error of mean (sem) across participants were plotted for depressed mood, as well as for primary and off target domain changes in intervention phase 2 relative to digital monitoring phase 1.

## Supplementary Results

**Supplementary Table 1** shows the full demographic breakdown of the N=50 participants.

**Supplementary Table 1:** Demographics for n=50 trial participants. sd: standard deviation.

| Demographics |  |
| --- | --- |
| Age (years, mean ± sd) | 43.3 ± 16.1 |
| Gender n (%) |  |
| Male | 12 (24%) |
| Female | 38 (76%) |
| Ethnicity n (%) |  |
| Caucasian | 32 (64%) |
| Black/African American | 5 (10%) |
| Asian | 6 (12%) |
| American Indian/Alaska Native | 1(2%) |
| More than one race | 4 (8%) |
| Unknown | 2 (4%) |
| Pre-intervention Mental Health (mean ± sd) | |
| PHQ9 | 9.2±5.1 |
| GAD7 | 7.1±5.3 |

Automated iMAP assignment. Here, we investigated whether a decision algorithm (DA) or an LLM (Google Gemini) could be used for iMAP target lifestyle domain assignment that resembled the selection made by the human coach. Supplementary Figure 3 shows the percent of times these models ranked the iMAP assigned by the human coach as their top first or second ranked choice. The naïve DA model resembled the human coach-based target domain assignment in 87.5% of cases (35 of 40 participants). It generated lifestyle domain scores and ranked these based on the top features within a domain (i.e., sleep/exercise/diet/social) in the personalized ML Shapley results, and further considered the independent correlation between feature variables and depression, what ratio of actionable features versus total features within a domain appeared in the top 10 Shapley ranks, and whether participants had low satisfaction within that domain indicating need for intervention.

The Gemini LLM approach yielded 92.5% match with the human coach iMAP domain assignment; this prompt also considered the same factors as the naïve DA model above. Finally, we also fine-tuned the DA model by empirically iterating the weights of the naïve DA equation based on the actual target domain preferences made by the human coaches across all study participants. The fine-tuned DA equation achieved 95% match with the human coach-based lifestyle domain assignment.

**Supplementary Figure 3.**
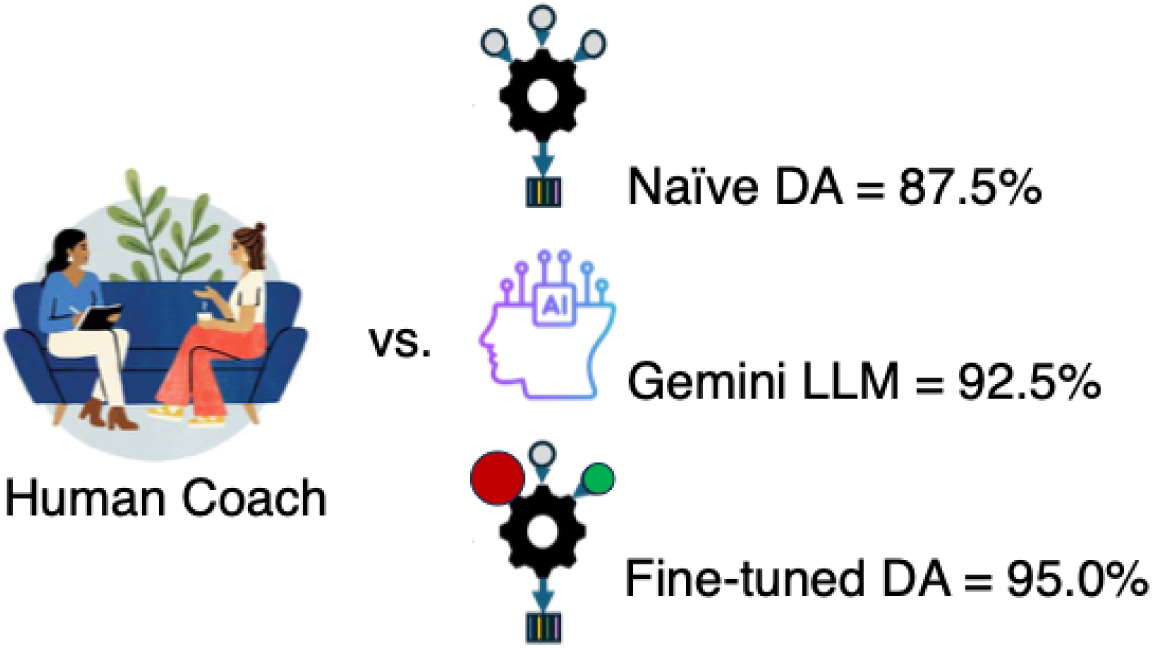
Representation of percent match between automated iMAP target lifestyle domain assignment versus human coach based assignment. DA: decision algorithm, LLM: large language model.

To further elucidate the separation between domains, we present the fine tuned DA scores for each participant shown in Supplementary Table 2.

**Supplementary Table 2:**
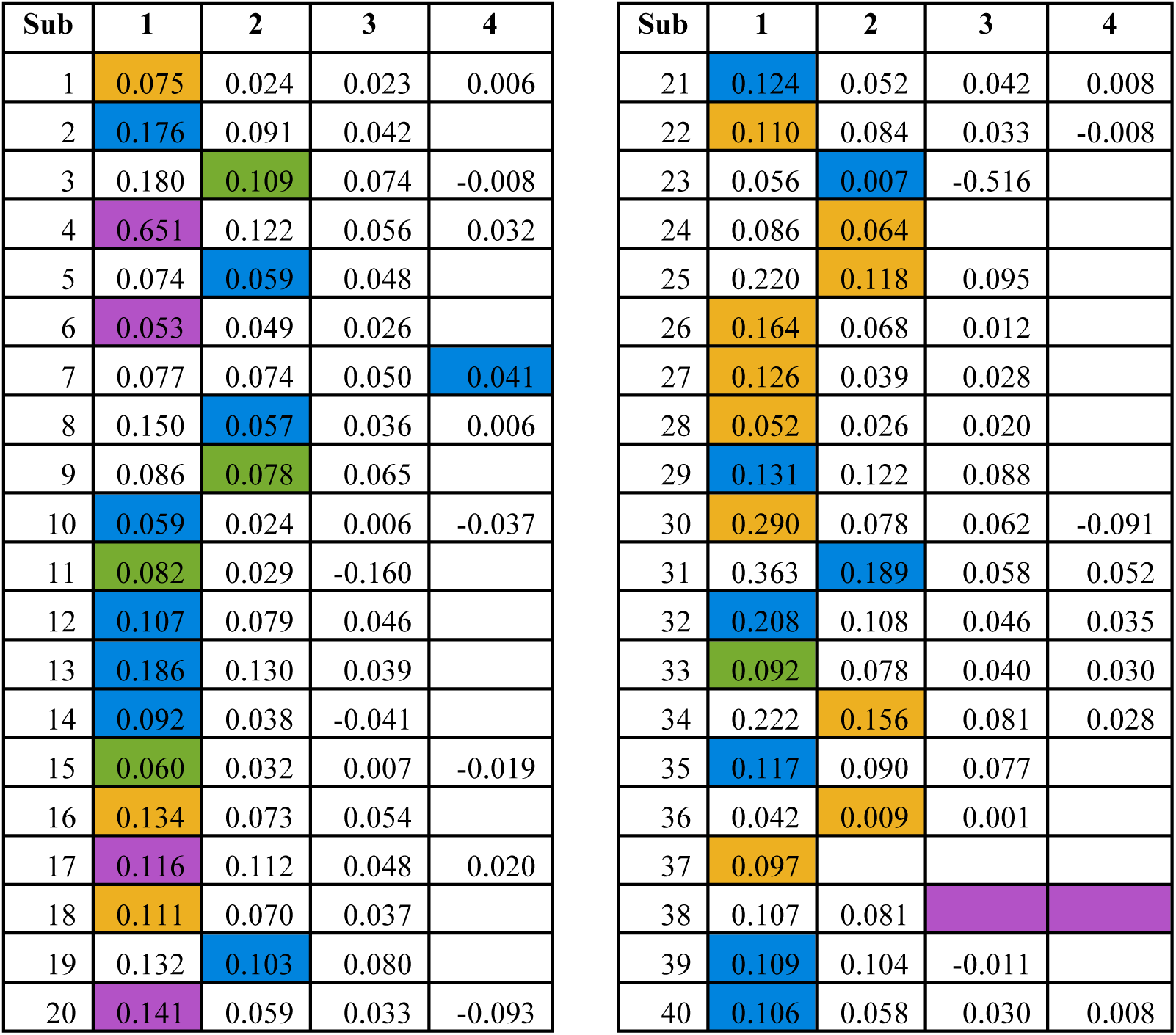
Fine-tuned algorithm domain rank list and scores per eq. 2 above for each participant. Domain scores are ordered from highest to lowest. Colored cells denote the iMAP domain assignment made by the human coach that was matched by the decision algorithm in the first or second rank for 38 of 40 participants. Blank cells indicate no score if domain variables were not within the top 10 ranked Shapley variables. Purple: Sleep; Orange: Exercise; Green: Diet; Blue: Social Connection.

**Supplemental Figure 4** shows our primary result (PHQ9) across the study period stratified across four assigned iMAPs.

**Supplemental Figure 4.**
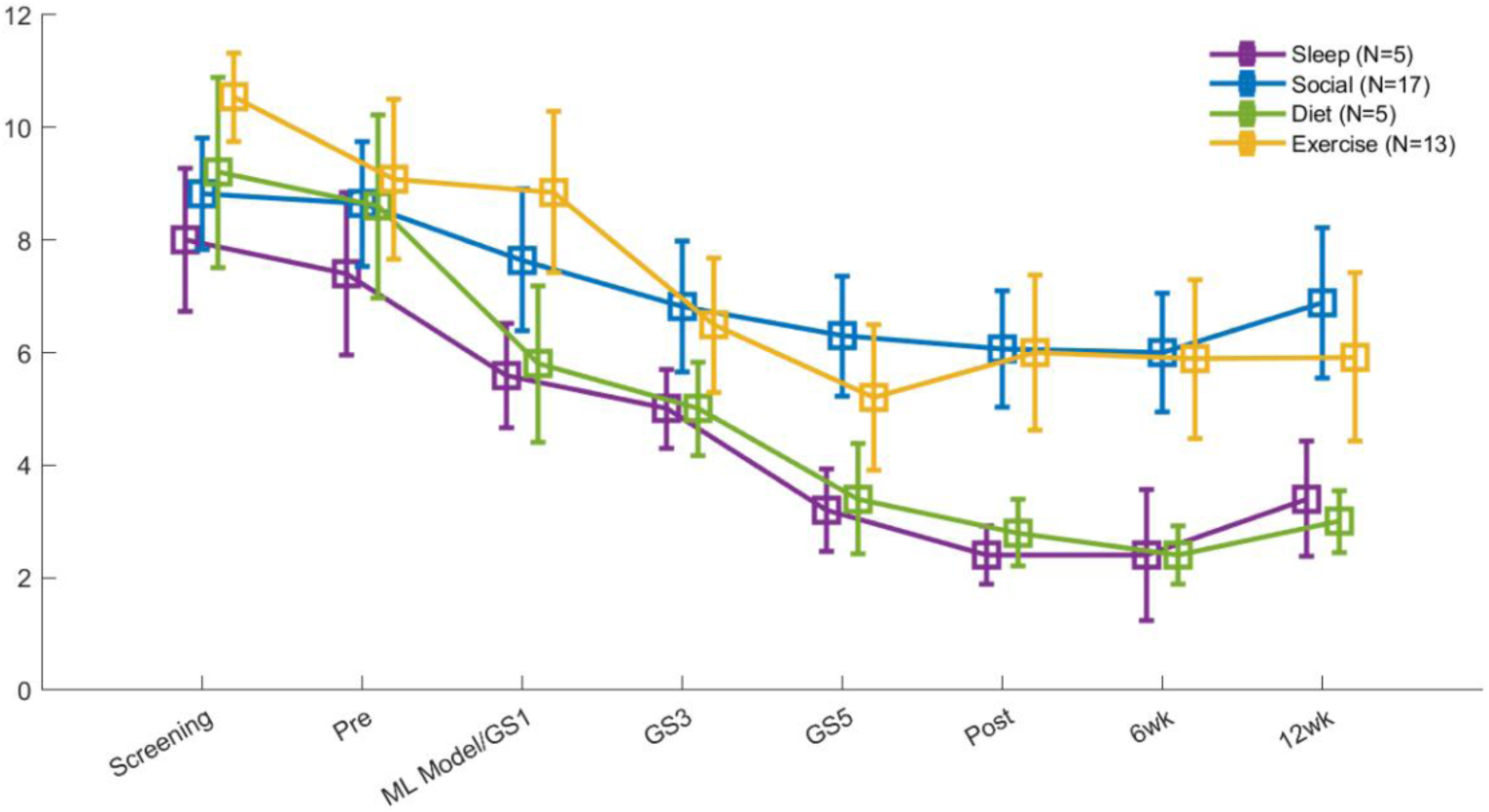
PHQ9 depression scores during the study period split by the four assigned iMAP domains. Mean ± standard error (sem) values are shown.

Relationship between primary and secondary outcomes. We investigated the relationship between post vs. pre-intervention change in the primary PHQ9 outcome and all secondary outcomes controlling for demographics using robust linear regression. This overall model was significant (F=3.5, p=0.003) and the only variable that showed a significant effect was quality of life (MCS12: β=-0.59±0.14, p=0.0004), i.e., improved quality of life was related to lower depression symptoms at post-intervention (Supplementary Figure 5A). Further, robust regression models at 6 and 12- week follow-ups were also significant, i.e., post vs. pre change in quality of life also predicted sustained improvement in PHQ9 scores from baseline to 6-week (β=-0.71±0.15, p=0.0002) and 12-week (β=-0.59±0.15, p=0.002) follow-ups.

Measures associated with remission. We used ANOVAs to investigate whether any pre-intervention data, i.e. demographics or secondary behavioral or cognitive outcomes showed separation between remitted vs. non-remitted trial participants (post-PHQ9 score<5). This analysis revealed that mindfulness (MAAS: F(df=1)=9.04; p=0.04) and anxiety (GAD7: F(df=1)=11.5; p=0.03) at baseline were significantly different between remitters (22/40) vs. non-remitters (18/40), with remitters showing lower anxiety and greater mindfulness at baseline (Supplementary Figure 5B). Of note, baseline anxiety and mindfulness were still significant predictors of remission (p<0.05) if baseline depression was accounted for in a generalized linear regression model. A similar analysis for intervention responders (≥ 50% symptom reduction, 16/40) vs. non-responders (24/40) showed no significant relationship after fdr-correction.

**Supplemental Figure 5.**
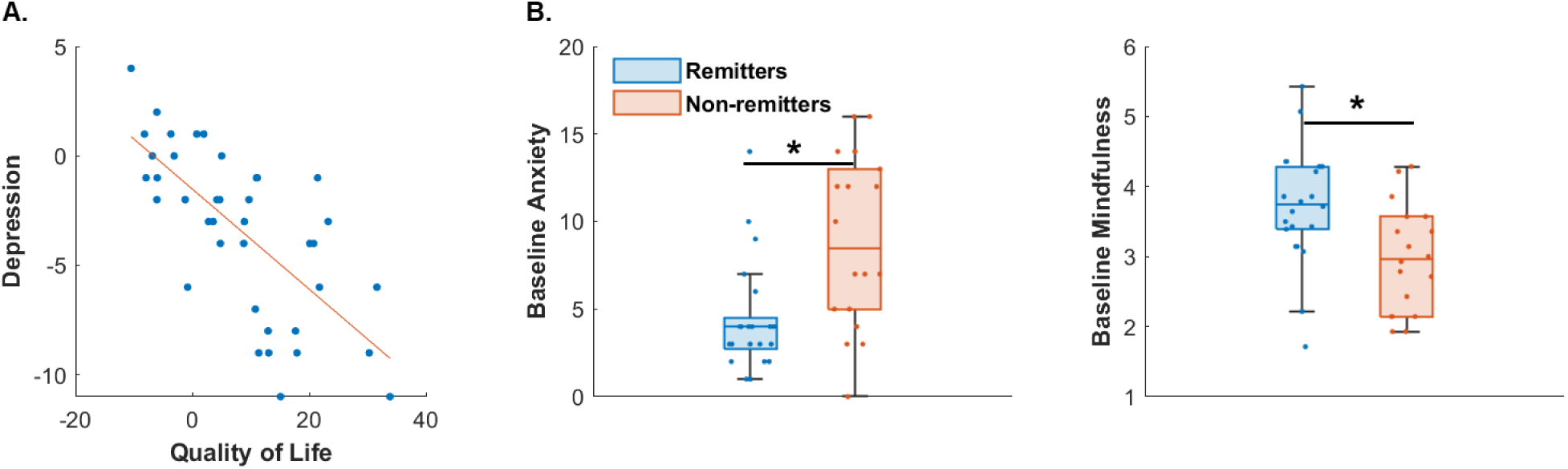
Factors supporting depressive symptom improvement. (A) Change in post vs. pre PHQ9 depression symptoms were associated with improvement in quality of life; scatter plot are shown with best fit line. (B) Baseline, i.e., pre-intervention measures associated with remission included anxiety (GAD7, left) and mindfulness (MAAS, right). Box plots show median with lower and upper quartiles as the box edges, whiskers denote the data range (excluding outliers) and the scatter points show individual participant values. Remitters depicted in blue and non-remitters in red. *p<0.05 fdr-corrected

## Supplementary Discussion

In our supplementary analysis, we developed a streamlined system using a combination of LLMs and algorithms that can automatically and accurately assign iMAPs. Our additional analysis also harmonizes our primary and secondary outcomes, and introduces a potential baseline marker for remission.

Our main approaches for automated iMAP assignment achieved an 87.5% (DA) and 92.5% (LLM) match with the coach’s decision, demonstrating that in a majority of cases iMAP assignment can be a straightforward process. The DA approach is transparent in that it is a simple structured equation that outputs a domain rank list for the preferred iMAP for each individual. LLM has the advantage of providing a logical explanation that may assist the coach in relaying personalized insights to the participant, yet the underlying model is a blackbox. Hence, the two approaches are complementary. Both the naïve DA and the LLM were designed a priori, i.e., with no iterations with regard to the actual iMAP assignments in the study to avoid overfitting to the dataset. With further empirical fine-tuning of the DA based on the actual coach iMAP assignments, the algorithmic match was improved to 95%. Yet the empirically-determined fine-tuned DA weights are overfit to our sample data and suitable only as a proof of concept. The 5% error rate of the fine-tuned DA may also be an underestimate given our modest sample size, hence, full automation without any human coach review is not yet recommended. Nevertheless, the complementary DA + LLM approaches may be useful assists for a human coach in future work.

In addition to our main findings, we also found that improvements in quality of life (QOL), i.e., how participants felt about their psychosocial functioning and ability to derive pleasure from their life activities ^65^, was significantly related to improvement in depression at post-intervention as well as at follow-ups. This is aligned with evidence showing that improvements in all lifestyle domains that we targeted is associated with improvements in the QOL mental component summary measure ^6,7,22,66,67^. Further, QOL improvement has been evidenced to predict lower odds of depression relapse and recurrence and thus, is a very important outcome for interventions ^68^.

Lastly, we found that individuals with greater mindfulness and lower anxiety scores at baseline were more likely to remit post-intervention, and these associations were significant even when controlling for baseline depression scores. Both of these factors have been linked to self-efficacy ^69,70^, i.e., belief in one’s ability to make change. Mindfulness has also been shown to facilitate behavior change through greater self-awareness and control over decision-making ^12,14^, which is why brief mindful attention to breathing was part of the digital schedule of the study ^10^. Overall, this result highlights the importance of greater self-efficacy, via greater mindfulness and lower anxiety to effect depression remission.

